# Extended-spectrum beta-lactamases resistance in *Klebsiella pneumoniae* in sub-Saharan Africa: a systematic review and meta-analysis

**DOI:** 10.1101/2024.08.10.24311782

**Authors:** Morufat Oluwatosin Olaitan, Oluwatosin Qawiyy Orababa, Bushola Rukayya Shittu, Adams Alabi Oyediran, Gift Maureen Obunukwu, Margaret Toluwalayo Arowolo, Ayomikun Emmanuel Kade, Khalid Ibrahim Yahaya, Rildwan Alaba Yusuff

**Affiliations:** Department of Biology, Microbiology and Science Laboratory Technology, Nile University of Nigeria, Abuja, Nigeria; School of Life Sciences, University of Warwick, Coventry, United Kingdom; Department of Microbiology, Osun State University, Osogbo, Nigeria; Department of Microbiology, University of Ibadan, Ibadan, Nigeria; Department of Microbiology, University of Lagos, Akoka, Lagos, Nigeria; Department of Microbiology, Federal University of Dutsin-Ma, Katsina, Nigeria; Department of Microbiology, University of Ilorin, Nigeria

## Abstract

**Background:** Extended-spectrum beta-lactamase (ESBL)-producing *Klebsiella pneumoniae* is a critical priority pathogen for which there is a need for new antimicrobials and poses a great public health threat to many parts of the world including sub-Saharan Africa (SSA). This study aims to determine the prevalence of ESBL-resistant *K. pneumoniae* in SSA and the predominant ESBL genes in the region.

**Methods:** Databases such as PubMed, Scopus, Web of Science, Africa Journal Online, and Google Scholar were searched for eligible articles based on preset eligibility criteria. After screening of titles, abstracts, and full texts, a meta-analysis using a random-effect model was conducted on the eligible studies to determine the overall and subgroup prevalence of ESBL-producing *K. pneumoniae* in SSA.

**Findings:** This meta-analysis included 119 eligible studies from 25 SSA countries in all SSA subregions. The overall prevalence of ESBL-resistant *K. pneumoniae* in SSA is estimated to be 8·6% [95% CI: 6·4-11]. South Africa (18·5%) and Central Africa (4·6%) subregions have the highest and lowest prevalence of ESBL-producing *K. pneumoniae* in the region, respectively. Additionally, South Africa (23·3%), Kenya (23%), and Nigeria (11·1%) are countries with the top three prevalence of ESBL-resistant *K. pneumoniae* in the region. Animal samples were also seen to have the highest prevalence compared to clinical and environmental samples in this study. Lastly, CTX-M-15 was the most reported ESBL gene in SSA.

**Interpretation:** Although this study reports a low pooled prevalence of ESBL-resistant *K. pneumoniae* in SSA, some countries in the region have a high burden of this drug-resistant strain. Additionally, some countries in the region lack data on this drug-resistant strain, thus putting other parts of the region at risk due to the porous borders and immigration between the countries in the region.

**Funding:** There was no funding for this study

## Introduction

Increased treatment failure of infectious diseases caused by microbes globally is attributed to antimicrobial resistance (AMR), which occurs due to widespread or indiscriminate use of antibiotics. AMR significantly impacts mortality and morbidity, bringing substantial economic burdens to people and countries.^1^ An estimation revealed five million AMR-linked deaths in 2019, which portend that the projected ten million deaths in 2050 and 24 million people below the poverty lines in 2030 as a result of AMR may be attained sooner than initially anticipated.^2,3^ Most of the deaths were believed to occur in sub-Saharan Africa and were attributed to infections caused by a cohort of both gram-positive and negative bacteria called ESKAPE pathogens. The ESKAPE pathogens include *Escherichia coli*, *Staphylococcus aureus*, *Klebsiella pneumoniae*, *Acinetobacter baumannii, Pseudomonas aeruginosa*, and *Enterococci faecium.*^4^

*Klebsiella pneumoniae,* a widely distributed Gram-negative, encapsulated bacterium, is a ubiquitous presence in our environment, found in water, soil, and the gut of healthy humans and animals.^5^ Classified under the family *Enterobacteriaceae*, they are known to be non-motile and lactose fermenting. *K. pneumoniae* is notorious for its role as an opportunistic pathogen, capable of causing both community and healthcare-associated infections.^6^ In fact, ten percent of nosocomial bacterial infections are attributed to *K. pneumoniae*.^7^ These strains of *K. pneumoniae* have been further classified into classical, hypervirulent, and multi-drug resistant strains based on phenotypic and genotypic differences.^8^

*K. pneumoniae* is responsible for a range of difficult-to-treat infections, including pneumonia, sepsis, bloodstream infections, meningitis, pyogenic liver abscesses, urinary tract infections (UTIs), and wounds.^8^ They are particularly associated with ventilator-associated pneumoniae (VAP), and are the leading cause of bacteraemia among the Gram-negative members of the ESKAPE pathogens, with a relatively high mortality rate.^9–11^

A significant concern for *K. pneumoniae* has been the rate at which they develop antibiotic resistance, as they harbour a lot of AMR plasmids.^10,13^ Thus, *K. pneumoniae* is listed as a critical pathogen in need of urgent drug development by the World Health Organisation (WHO).^14^ Several studies have revealed the vast array of mechanisms adopted by this pathogen to evade both host immune defence and antibiotics, including the production of antibiotic-modifying enzymes such as Extended Spectrum Beta-Lactamase (ESBL).^15,16^ ESBL confers resistance to a group of antibiotics with a beta-lactam ring, including penicillin, monobactams (aztreonam), and the first, second, and third-generation cephalosporin by hydrolysing the ring.^6^ *K. pneumoniae* has been reported to harbour several plasmid-encoded ESBL enzyme families and their variants, such as Sulfhydryl variable (SHV), Temoniera (TEM), and Oxacillinases (OXA).^17^ In recent years, cefotaximase (CTX-M) has emerged, which is increasingly reported on a global scale and is currently the most predominant ESBL enzyme in numerous continents and countries worldwide.^18–20^

Despite growing concerns about ESBL-resistant *K. pneumoniae*, to the best of our knowledge, there are no available study on systematic review and meta-analyses of the pooled prevalence status in sub-Saharan Africa. Understanding the prevalence of these resistant strains is essential for guiding clinical decision-making, informing public health interventions, and ultimately improving patient outcomes in the face of growing antibiotic resistance. This systematic review and meta-analysis aim to comprehensively determine the prevalence of ESBL-producing *K. pneumoniae* in sub-Saharan Africa, describe the epidemiology within the region, and investigate the predominant ESBL genes in the region.

## Methods

### Study design, Search strategy and screening

This study is a systematic review and meta-analysis and was conducted in compliance with the Preferred Reporting Items for Systematic Reviews and Meta-Analysis (PRISMA) 2020 guideline for systematic reviews and meta-analysis.^21^ Databases such as PubMed, Scopus, Web of Science, Google Scholar, and African Journal Online were searched using combinations of the following keywords “ESBL”, “Extended-spectrum beta-lactamase”, “resistance”, “*Klebsiella pneumoniae*”, “*K. pneumoniae*”, “sub-Saharan Africa”, “SSA”. The last search was done in December 2023. The articles’ titles, abstracts, and full texts were screened to check if they comply with the preset eligibility criteria (Supplementary criteria).

### Data extraction and quality assessment

A data extraction table was created to include important details from the articles such as first author, year of publication, study design, sample source, country of study, sub-region, sample size, number of *K. pneumoniae* isolates, number of ESBL strains, and ESBL gene. All human samples were grouped as clinical sources in this study while samples from either food or water sources were put under environmental sources. The qualities of the studies were assessed based on the method used to characterise extended-spectrum beta-lactam strains. Studies that used only phenotypic methods like disc diffusion, broth microdilution, and automated antimicrobial susceptibility testing methods like VITEK were grouped as low, studies that used only genotypic/molecular characterisation methods were classified as medium, while studies that used both phenotypic and molecular methods were classified as high quality (Supplementary Figure 1).

### Meta-analyses

The pooled prevalence of extended-spectrum beta-lactamase-producing *K. pneumoniae* was estimated from the sample size and number ESBL isolates reported in the study using meta package in R (version 4.3.3). Subgroup analysis based on subregion, sample source, and country was done but only subgroups with more than two eligible articles are reported in this study. The sub-Saharan Africa map was created in R using the rnaturalearth package. Random effect model was used in this study due to the variations expected between the studies which includes sample size and study settings. Funnel plot of the overall effect size was used to visualise publication bias.

## Results

### Search outcome

A total of 4746 search results were retrieved from four databases namely Google Scholar (4230), PubMed (162), Scopus (270), Web of Science (28), and African Journal Online (56). With Google Scholar, only the first five pages were considered. One hundred and nineteen (119) eligible studies were finally included in the meta-analysis after the screening of titles, abstracts, full texts, and the exclusion of duplicates and ineligible studies (Figure 1).

**Figure 1.**
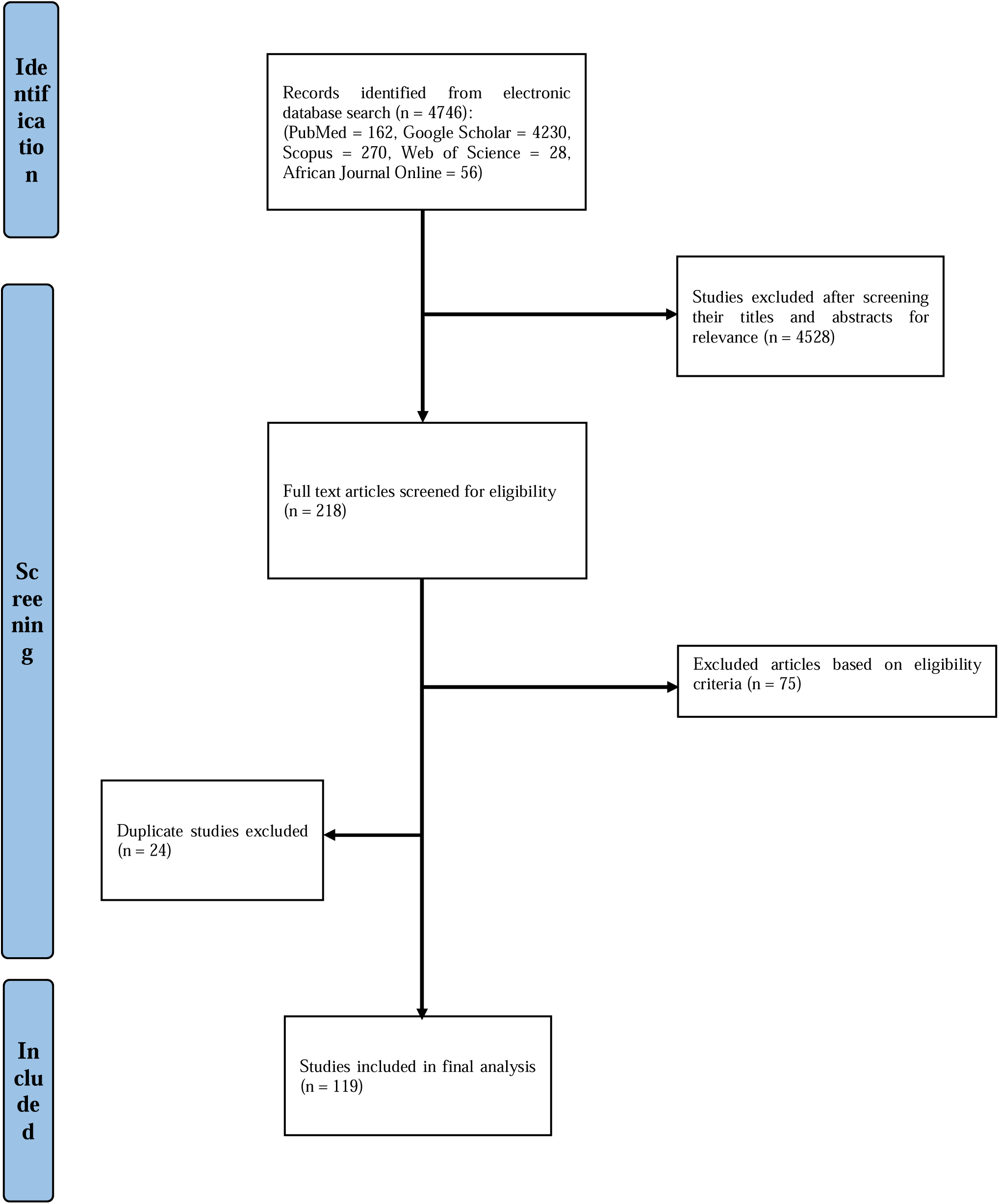
PRISMA Flow diagram of search and screening strategy.

### Study characteristics

This study analysed a total of 119 eligible articles from 25 sub-Saharan African countries. East Africa (53) had the highest number of studies reporting ESBL resistance in *K. pneumoniae*, followed by West Africa (43), South Africa (12), and Central Africa (10) (Figure 3). One of the articles reported ESBL resistance in *K. pneumoniae* from multiple sub-regions in SSA. Additionally, Ethiopia (20) had the highest number of eligible studies, followed by Tanzania (16), Nigeria (14), Ghana (12), South Africa (ten), Uganda (six), Gabon (four), Malawi (three), Cameroon (four), Kenya (four), Madagascar (three), Benin (three), DRC (three), Sierra Leone (two), Chad (two), and Burkina Faso (two). Others (Mali, Zimbabwe, Gambia, Sudan, Togo, Guinea Bissau, Mozambique, Côte d’Ivoire, and Central Africa Republic) had just one eligible study each while one study was carried out in multiple countries. Most of the eligible articles reported ESBL resistance in *K. pneumoniae* from clinical samples (104), followed by environmental (ten), and animal (five) samples (Figure 4). The characteristics of the 119 eligible studies are presented in the Supplementary Table 1.

**Figure 2.**
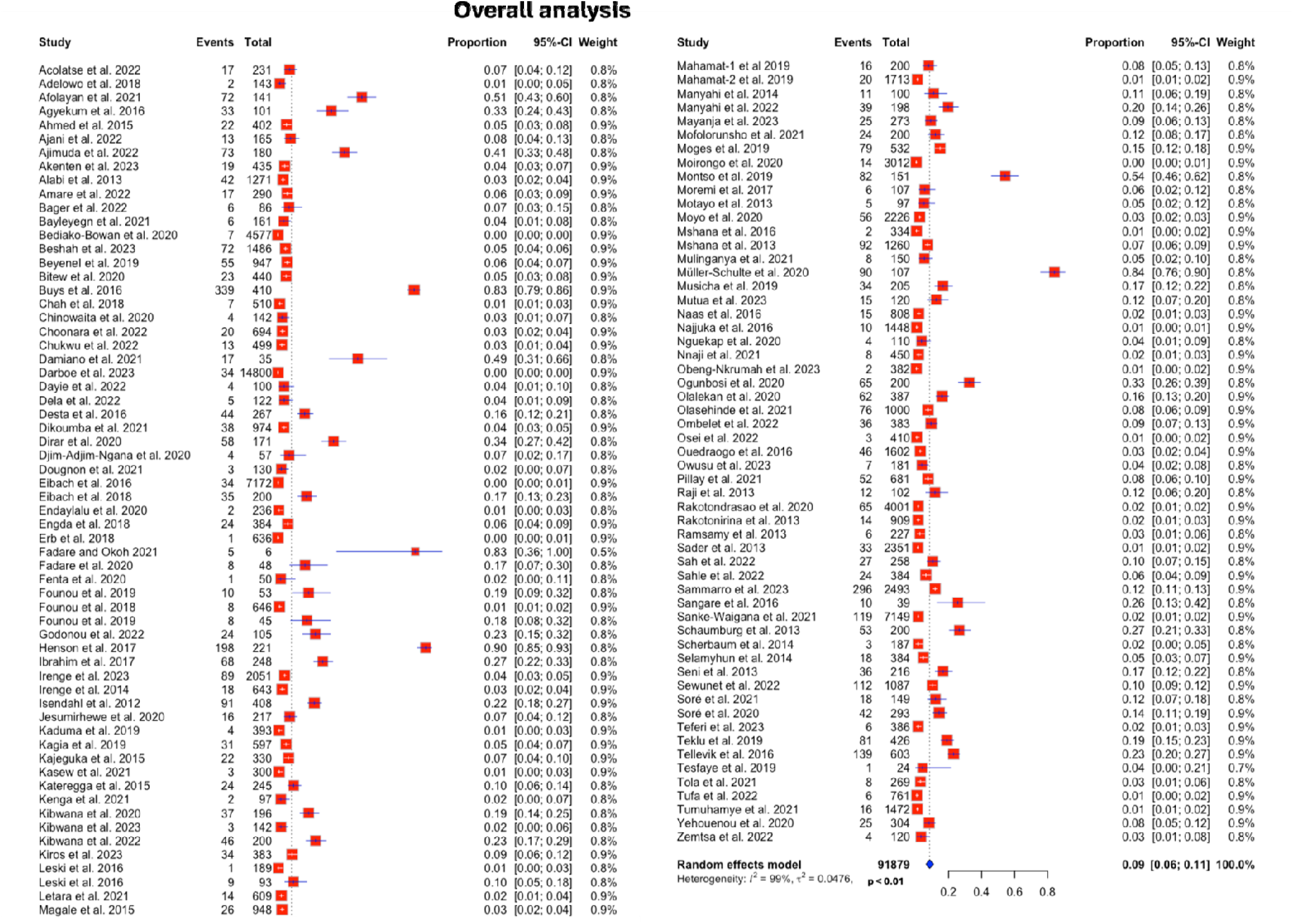
Forest plot of overall prevalence of ESBL-producing *K. pneumoniae* from sub-Saharan Africa.

**Figure 3.**
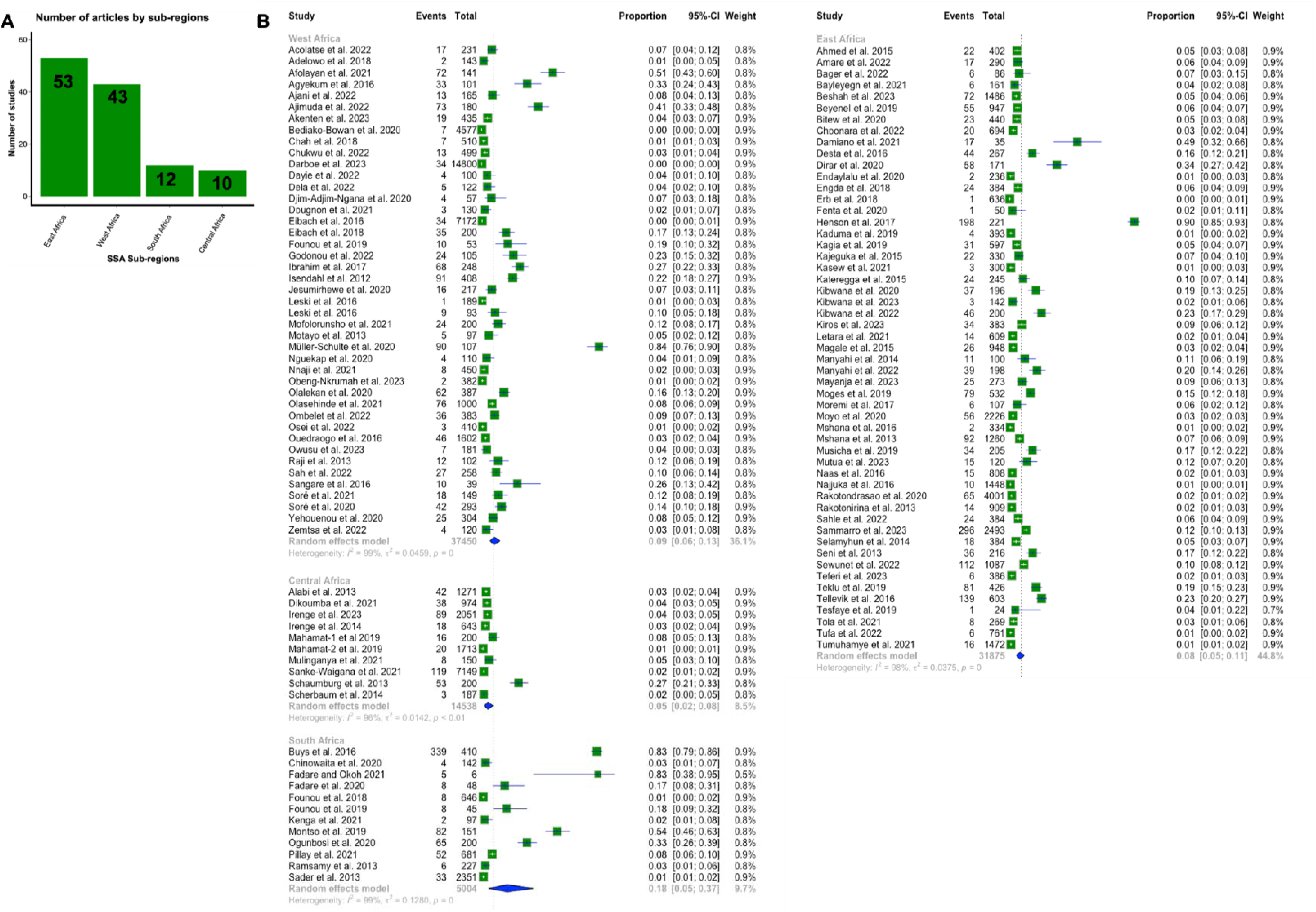
Forest plot showing subgroup analysis of ESBL-producing *K. pneumoniae* based on subregions in sub-Saharan Africa.

**Figure 4.**
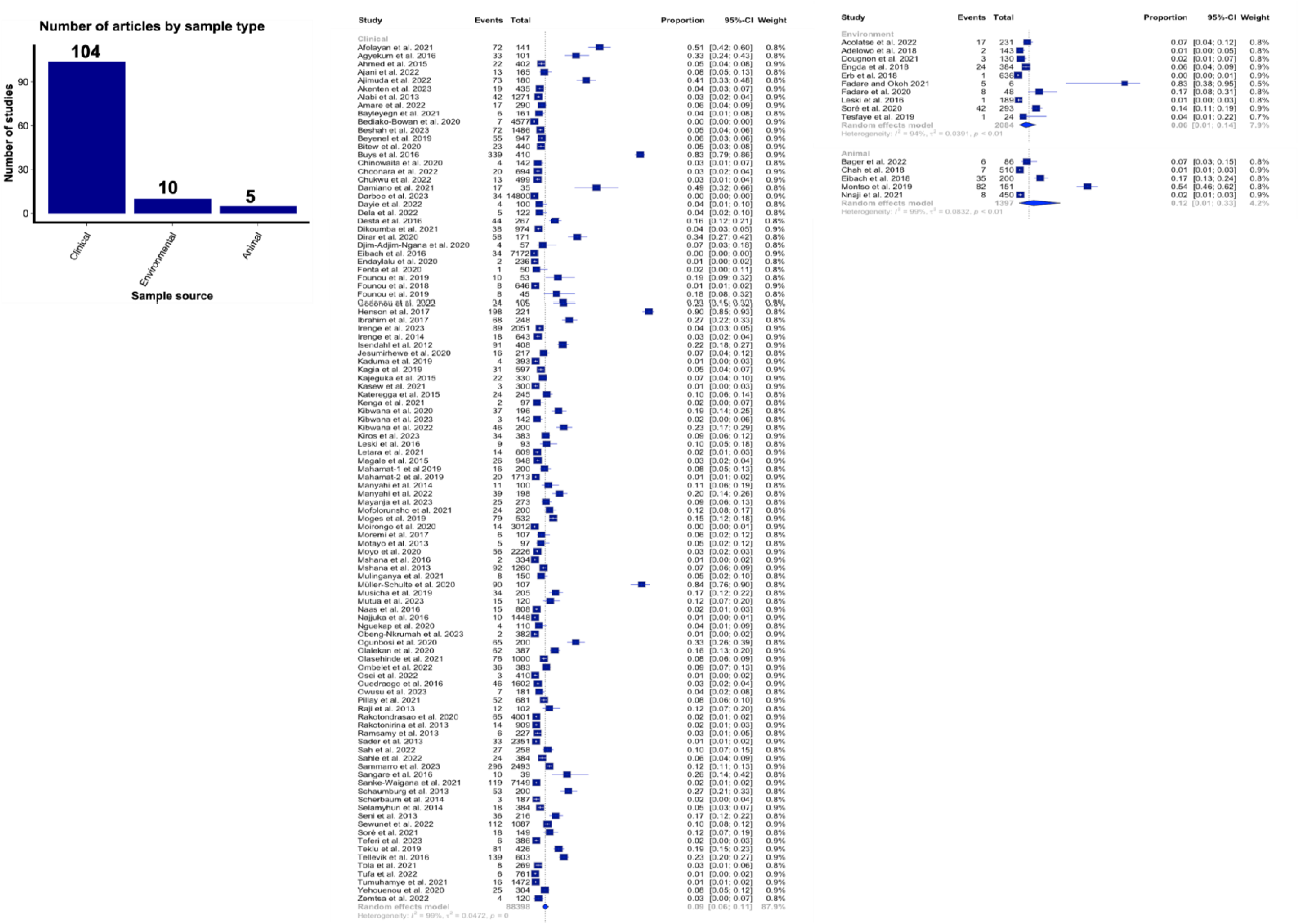
Forest plot showing subgroup analysis of ESBL-producing *K. pneumoniae* based on sample source in sub-Saharan Africa.

**Figure 5.**
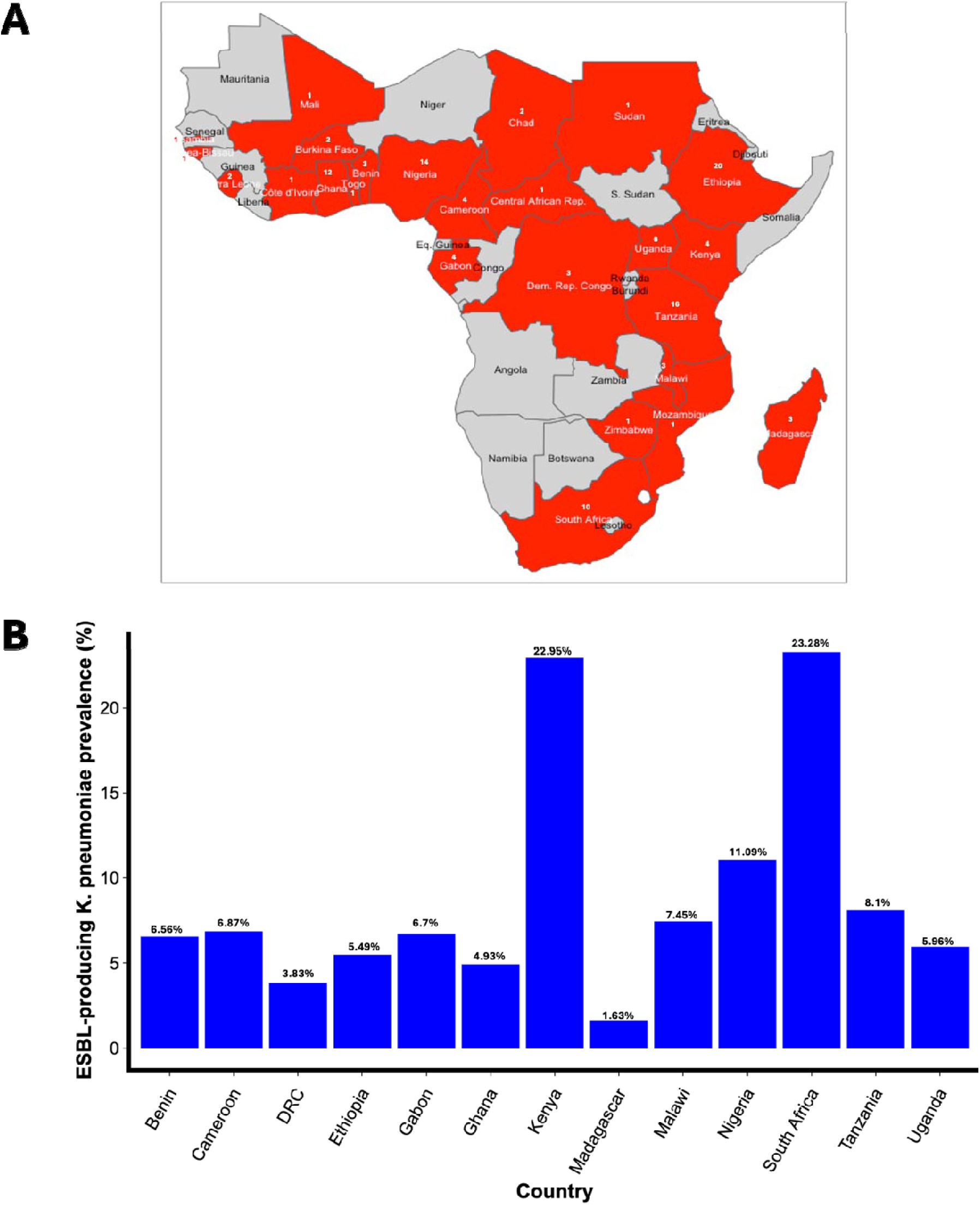
A. Map of sub-Saharan Africa highlighting countries with eligible articles (in red) B. The estimated country prevalence of ESBL resistance in K. pneumoniae with their corresponding prevalence.

### Prevalence of ESBL-resistance in *K. pneumoniae*

The pooled prevalence of ESBL-resistance in *K. pneumoniae* in sub-Saharan Africa is estimated to be approximately 8·6% [95% CI: 6·4-11·0] (Figure 2). The South African subregion was estimated to have the highest prevalence (18·5% [95% CI: 5·07-37·1]) of ESBL-producing *K. pneumoniae*, followed by West Africa (9·3% [95% CI: 5·8-13·4]) (Figure 3). On the other hand, the subregion with the lowest prevalence was Central Africa (4·6% [95% CI 1·9-8·3]) (Figure 3). The highest ESBL prevalence in *K. pneumoniae* in SSA was seen in animals (12·1% [95% CI: 0·8-33·1]) compared to 8·6% [95% CI: 6·4-11·2]) from clinical specimens and 6·2% [95% CI: 0·97-14·4] from the environment (Figure 3). Among the countries included in this analysis (countries with at least three eligible studies), South Africa (23·3% [95% CI: 6·5-45·8]) has the highest burden of ESBL-resistant *K. pneumoniae*, followed by Kenya (23% [95% CI: 0-68]), Nigeria (11·1% [95% CI: 5·1-18·9]), and Tanzania (8.1·% [95% CI: 3·8-13·8]) (Figure 4). Madagascar had the lowest prevalence of ESBL-resistant *K. pneumoniae* (Figure 4). The forest plot of subgroup analysis of ESBL-producing *K. pneumoniae* based on countries in sub-Saharan Africa is presented in the Supplementary Figure 2. A high *I*^2^ value was seen in this result which is an indication of high heterogeneity in this study. Funnel plot was also asymmetrical (Supplementary Figure 3).

### Genomic epidemiology of ESBL genes in SSA

Fifty (50) out of the 119 eligible studies included in this analysis used genotypic assays to characterise ESBL genes in *K. pneumoniae* (Supplementary Figure 1). The ESBL genes reported in the studies include CTX-M (including CTX-M-1, CTX-M-2, CTX-M-8, CTX-M-9, CTX-M-11, CTX-M-14, and CTX-M-15), SHV (SHV-11, SHV-12, SHV-28), TEM, and OXA. Among these genes, CTX-M-15 was the most reported in most articles analysed. Additionally, all three major ESBL genes (CTX-M, SHV, and TEM) were reported in all the sample types (clinical, animal, and environment).

## Discussion

*Klebsiella pneumoniae* remains an important human and animal pathogen due to its position as a member of the ESKAPE pathogens and a WHO’s critical priority pathogen. Additionally, the presence of ESBL genes in *K. pneumoniae* confers resistance to many antibiotics, hence, making it difficult to treat infections resulting from these resistant strains. Understanding the prevalence of ESBL-resistant *K. pneumoniae* strains in sub-Saharan Africa is an important step to curbing the spread of this pathogen within the region as well as to other parts of the world.

This study analysed articles that reported ESBL-resistance in *K. pneumoniae* from clinical, environmental, and animal sources across 25 countries in sub-Saharan Africa (SSA). The overall prevalence of ESBL-resistant *K. pneumoniae* in sub-Saharan Africa is 8·6%. Similarly, the highest prevalence of ESBL-producing *K. pneumoniae* in SSA subregion was estimated at 18·5% in Southern Africa, followed by 9·3% in West Africa, and the lowest at 4·6% in Central Africa. These are lower than what has previously been reported in South East Asia (27%), Middle East (35·4%),^22^ and elsewhere.^23^ Paradoxically, the recent global burden of AMR report estimated sub-Saharan Africa as one of the highest burdens of AMR.^2^ However, this low prevalence might have cropped up from the point that many laboratories and healthcare facilities in the region lack familiarity with the significance of detecting ESBL-producing Gram-negative organisms. This might have contributed to the low prevalence. Notwithstanding, Mohd et al.^23^ reported a 28·7% prevalence of MDR *K. pneumoniae* in Africa which is a much higher value than we reported in this study. The disparity might be due to the fact that the authors considered all countries in Africa while ours only focused on SSA. Moreover, the authors considered all multidrug-resistant strains of *K. pneumoniae*, including carbapenem-resistant strains, whilst ESBL-resistant strains are our focal point.

Animal studies had the highest prevalence (12·1%) followed by clinical studies (8·6%) and environmental studies (6·2%). Since their introduction in the 1940s, antimicrobials have significantly impacted human and animal health. Over the decades, these drugs have been extensively used in animals for disease control, prevention, and treatment, as well as growth promotion in animal husbandry. ^24^ Antimicrobial growth promoters (AGPs) have been employed in the United States and other developed countries for over 50 years, with their introduction dating back to the mid-1950s. Moore and Stokstad first documented the benefits of AGPs (sulfasuxidine, streptothricin, and streptomycin) in chicken and pig feed, noting improvements in animal production.^25^ Subsequent studies have confirmed similar benefits from other antimicrobials in food animal production. In the US, sub-therapeutic antimicrobial use in food animals reached approximately 14600 tons in 2012.^26^ China, the leading producer and consumer of antimicrobials globally, used 29774·09 tons for animal husbandry in 2018, with 53·20% of this amount allocated for growth promotion.^27^

Antimicrobials have also been found in fish and other aquaculture products. For instance, chloramphenicol was detected in carp and chub muscle at levels exceeding permissible limits.^28^ A separate study identified norfloxacin, ciprofloxacin, and enrofloxacin in nine marine fish species.^29^ Prolonged antibiotic use in food animals fosters the development and spread of resistant strains. These resistant bacteria can reach humans directly or indirectly through food, water, mud, and manure used as fertilizers. Evidence shows that resistant bacteria are present in foods from animal sources and throughout food processing stages.^30^ Horizontal gene transfer (HGT) between bacterial species, facilitated by mobile genetic elements like plasmids and transposases, significantly contributes to the rapid spread of resistance.^31^ Those in close contact with infected animals, such as farm and slaughterhouse workers and veterinarians, are at higher risk of exposure to resistant bacteria.

Extended use of subtherapeutic antimicrobial doses in animal production creates ideal conditions for bacteria to thrive and develop resistance genes, which can then be transmitted to human gut microbiota through contaminated food or environmental exposure.^30,32^ Research into the transmission dynamics of resistant pathogens enhances our understanding of how these pathogens spread among humans.^33^ Enterobacteriaceae, including opportunistic pathogens like *Escherichia coli* and *Klebsiella* spp, are commonly found in livestock and on retail meat, often causing urinary tract and bloodstream infections in patients.^34^

Additionally, ESBL-producers are resistant not only to beta-lactams but also to other antibiotics like tetracyclines, fluoroquinolones, aminoglycosides, and trimethoprim/sulfamethoxazole.^35^ Factors contributing to this high level of resistance include improper use of antimicrobials by healthcare professionals and self-prescription by patients.^36^ To address this issue, surveillance systems should adopt a ‘One Health’ approach, which integrates efforts across human, animal, and environmental health. Reports indicate a growing presence of antibiotic-resistant Gram-negative bacteria, particularly Enterobacteriaceae, in hospital wastewater.^37^ The widespread use of antibiotics in animals, which are structurally similar to those used in humans, exacerbates the problem of antibiotic-resistant bacteria affecting public health. Resistant bacteria and genetic elements can spread through feces, environmental contamination, and animal food sources, affecting all stages of food processing.^38^

Among countries with at least three eligible studies, South Africa had the highest burden of ESBL-resistant *K. pneumoniae* at 23·3%, followed by Kenya at 23%, Nigeria at 11·1%, and Tanzania at 8·1%. Madagascar reported the lowest prevalence (1·63%). The prevalence of ESBL-resistant *K. pneumoniae* in some sub-Saharan African countries in this study is similar to previous reports in Egypt.^39^ They reported prevalence rates of 17% and 38·8%, suggesting an increasing spread of ESBLs, possibly due to the widespread use of third-generation cephalosporins. Prevalence varies by species, region, infection control practices, and antibiotic use patterns. Selective pressure from excessive cephalosporin use in certain countries contributes to rising ESBL rates.^40^

This analysis shows that there are still countries in sub-Saharan Africa with no study on ESBL-resistant *K. pneumoniae*. These grey areas pose a great challenge in curbing the spread of AMR pathogens and genes. The ease of migration and porous borders between these countries and other countries within the region – especially those that share the same boundaries – would make it easy to spread multidrug-resistant pathogens like *K. pneumoniae*.^41^ To avoid this, both country and regional governments in SSA should deploy resources to grey areas to help understand the epidemiology of multidrug-resistant pathogens and control their spread. Additionally, some of the countries in this study (Côte d’Ivoire, Central Africa Republic, Zimbabwe, Mozambique, Togo, Gambia, Guinea Bissau, Mali, Burkina Faso, Chad, Sudan and Sierra Leone) have less than 3 eligible articles, making it difficult to estimate the prevalence of ESBL-resistant *K. pneumoniae* in these countries.

Globally, there is growing concern about how ESBL-producing bacteria affect the effectiveness of treatments for bacterial infections. This study identifies the major ESBL genes present in *K. pneumoniae* isolates in sub-Saharan Africa. Our study revealed TEM, SHV, and CTX-M as the three main ESBL genes in *K. pneumoniae* in SSA. The detection of these resistance genes in *K. pneumoniae* suggests the spread of bacteria producing ESBL enzymes, specifically bla-beta-lactamases. Additionally, other beta-lactamase genes identified include carbapenem-hydrolyzing beta-lactamases such as OXA-1, OXA-10, OXA-48, OXA-1-like, metallo-beta-lactamase gene IMP, class C beta-lactamases AmpC (which inactivates first- and second-generation cephalosporins, including cephamycins like cefoxitin and cefotetan, and third-generation cephalosporins like ceftazidime), LEN (which confers resistance to ampicillin, amoxicillin, carbenicillin, and ticarcillin but not extended-spectrum beta-lactams), OKP (which confers resistance to penicillins and early cephalosporins in *K. pneumoniae*), and CMY-2 (which confers broad-spectrum resistance to beta-lactam antimicrobials, including ceftriaxone and ceftiofur, as well as beta-lactamase inhibitors like clavulanic acid). The CTX-M gene emerged as the most common, followed by SHV and TEM, consistent with findings from other regions.^42^ Despite numerous studies, some areas in SSA still underreport ESBL cases, leaving prevalence unclear. It is evident that CTX-M, SHV, and TEM are the most commonly detected genes, highlighting the need for comprehensive monitoring and infection control measures.

Research has shown that TEM genes have been reported in vegetables from Finland^43^ and southern Thailand,^44^ while CTX-M is the most commonly detected gene in animals, humans, and the environment. Additionally, *K. pneumoniae* and *E. coli* from dogs and cats have been found to possess CTX-M-9 type genes.^45^ CTX-M enzymes are now the most prevalent ESBL types, likely due to their environmental origins.^46^ These enzymes are classified into five subgroups based on amino acid composition: CTX-M-1, CTX-M-2, CTX-M-8, CTX-M-9, and CTX-M-25.^47^ Our results underscore the importance of the “One Health” approach, as ESBL-producing *K. pneumoniae* was found in animals, clinical specimens, and the environment. The presence of CTX-M and SHV in ESBL-producing *K. pneumoniae* from these sources further emphasizes the need for a “One Health” perspective. Future research should adopt this approach to better understand the links between human, animal, and environmental health, particularly regarding antibiotic resistance in zoonotic diseases.

Detection of ESBL *K. pneumoniae* in the studies appraised revealed utilization of various methods, including phenotypic assays, genomic assays, Polymerase Chain Reaction (PCR), NMK-203 card on the Phoenix system, Disc Diffusion (Standard and Kirby-Bauer methods), Etest, Double Disc Diffusion, Vitek 2 Compact System, Double Disc Synergy Test (DDST), Whole Genome Sequencing, Broth Microdilution, and EUCAST Breakpoints. The use of diverse detection methods contributed to significant heterogeneity in our study. Whole genome sequencing remains an important method for identifying antibiotic-resistant genes, however, many laboratories in SSA do not have resources for this. Hence, result to phenotypic detection methods which could give false positive or negative results. As a result, many antibiotic-resistant pathogens are under-reported in the region.

In conclusion, our study highlights the urgent need for sustained surveillance in sub-Saharan Africa of multiple drug-resistant microorganisms, especially in locations where data is scarce or non-existent. The high prevalence of these resistant strains in countries such as Tanzania, Nigeria, Kenya, and South Africa is particularly alarming given the region’s porous borders and the potential for cross-border transmission. The underreporting of antimicrobial resistance in the region further exacerbates the threat, potentially allowing resistant bacteria to spread swiftly from areas with high, yet unreported, prevalence to those with lower documented cases. To effectively identify and mitigate these emerging threats, it is imperative that policymakers in sub-Saharan Africa prioritize the establishment of comprehensive surveillance systems to curb the spread of multidrug-resistant infections in the region.

## Data Availability

All dataset generated and analysed in this study are included within the article and / or its Supplementary data file.

## Authors’ Contribution

**Conceptualization**: Morufat Oluwatosin Olaitan

**Data curation**: Morufat Oluwatosin Olaitan, Oluwatosin Qawiyy Orababa, Bushola Rukayya Shittu, Adams Alabi Oyediran, Gift Maureen Obunukwu

**Formal analysis:** Oluwatosin Qawiyy Orababa

**Investigation:** Morufat Oluwatosin Olaitan, Oluwatosin Qawiyy Orababa, Bushola Rukayya Shittu, Adams Alabi Oyediran, Gift Maureen Obunukwu, Khalid Ibrahim Yahaya, Rildwan Alaba Yusuff

**Methodology**: Morufat Oluwatosin Olaitan, Oluwatosin Qawiyy Orababa, Bushola Rukayya Shittu, Adams Alabi Oyediran, Gift Maureen Obunukwu, Margaret Toluwalayo Arowolo, Ayomikun Emmanuel Kade, Khalid Ibrahim Yahaya, Rildwan Alaba Yusuff

**Project administration**: Morufat Oluwatosin Olaitan

**Resources**: Morufat Oluwatosin Olaitan, Oluwatosin Qawiyy Orababa

**Software**: Morufat Oluwatosin Olaitan, Oluwatosin Qawiyy Orababa

**Supervision**: Morufat Oluwatosin Olaitan, Oluwatosin Qawiyy Orababa

**Validation**: Morufat Oluwatosin Olaitan, Oluwatosin Qawiyy Orababa

**Visualization**: Morufat Oluwatosin Olaitan, Oluwatosin Qawiyy Orababa, Margaret Toluwalayo Arowolo

**Writing – original draft**: Morufat Oluwatosin Olaitan, Oluwatosin Qawiyy Orababa, Margaret Toluwalayo Arowolo, Ayomikun Emmanuel Kade

**Writing – review & editing**: Morufat Oluwatosin Olaitan, Oluwatosin Qawiyy Orababa

## Acknowledgments

None

