## Supplementary materials_Kp for "Extended-spectrum beta-lactamases resistance in *Klebsiella pneumoniae* in sub-Saharan Africa: a systematic review and meta-analysis"

Supplementary criteria …………………………………………………………........ 2

Supplementary Figure 1 ……………………………………………………………... 2

Supplementary Table 1 ………………………………………………………….…... 3

Supplementary Figure 2 ..……………………………………………………………18

Supplementary Figure 3 …………………………………………………………….... 19

Supplementary Material References of eligible articles included in this study ………..… 20

### Supplementary criteria

Articles that reported prevalence of ESBL-resistance *Klebsiella pneumoniae*, in sub-Saharan African countries, between 2013 and 2023, published in English, not a review or systematic review and/or meta-analysis.

### Qualities of studies

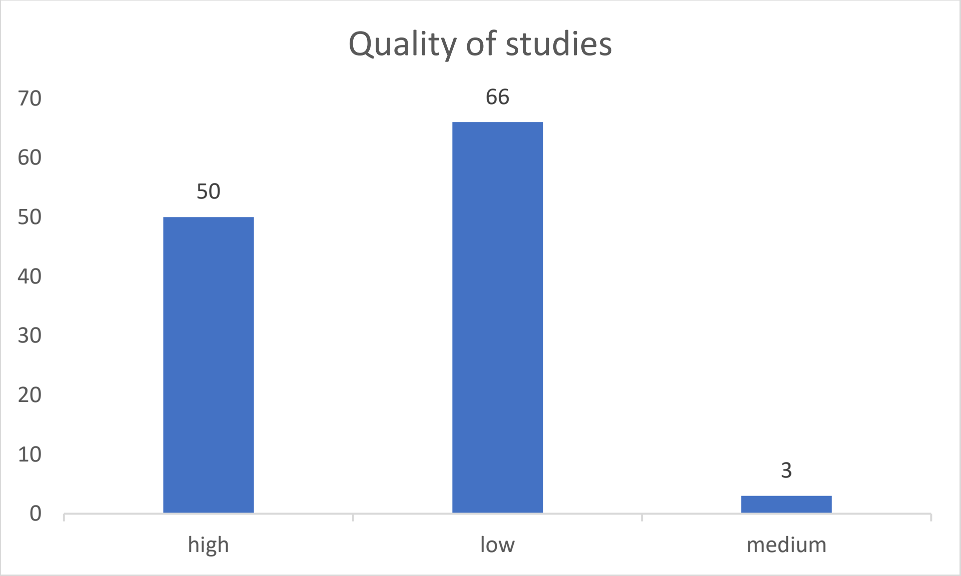


**Supplementary Figure 1**. Number of articles based on the quality of studies from sub-Saharan Africa. Research using only phenotypic techniques, such as broth microdilution, disc diffusion, and automated antimicrobial susceptibility testing techniques like VITEK, were categorized as low quality; research using only genotypic/molecular characterisation techniques was categorized as medium quality; and research using both phenotypic and molecular techniques was categorized as high quality.

**Supplementary Table 1. C**haracteristics of studies reporting ESBL-producing *K. pneumoniae* in sub-Saharan Africa

| **First author** | **Year** | **Study type** | **Country** | **Sub-region** | **Sample source** | **Sample size** | **Isolate number** | **ESBLprevalence** | **Method used** | **ESBL gene** | Quality |
|---|---|---|---|---|---|---|---|---|---|---|---|
| Acolatse et al.[1] | 2022 | Cross-sectional | Ghana | West Africa | Environment | 231 | 17 | 17 | Phenotypic and genotypic assay | SHV, CTX-M, TEM | high |
| Adelowo et al.[2] | 2018 | Cross-sectional | Nigeria | West Africa | Environment | 143 | - | 2 | Phenotypic and genotypic assay | SHV, CTX-M-15, TEM | high |
| Afolayan et al.[3] | 2021 | - | Nigeria | West Africa | Clinical | 141 | - | 72 | Molecular | CTX_M-15 | medium |
| Agyekum et al.[4] | 2016 | Cross-sectional | Ghana | West Africa | Clinical | 101 | 43 | 33 | NMIC-203 card on the Phoenix system | CTX-M-15, CTX-M | low |
| Ahmed et al.[5] | 2015 | Longitudinal | Tanzania | East Africa | Clinical | 402 | 20 | 22 | Disk diffusion | - | low |
| Ajani et al.[6] | 2022 | Cross-sectional | Nigeria | West Africa | Clinical | 165 | 26 | 13 | DDST, Etest and PCR | SHV, TEM | high |
| Ajimuda et al.[7] | 2022 | Cross-sectional | Nigeria | West Africa | Clinical | 180 | 172 | 73 | Phenotypic and genotypic assay | SHV, TEM, and CTX-M | high |

| | | | | | | | | | | | |
|---|---|---|---|---|---|---|---|---|---|---|---|
| Akenten et al.[8] | 2023 | Cross-sectional | Ghana | West Africa | Clinical | 435 | - | 19 | Phenotypic and genotypic assay | SHV, CTX-M, TEM | high |
| Alabi et al.[9] | 2013 | Retrospective study | Gabon | Central Africa | Clinical | 1271 | 48 | 42 | Disc diffusion, double-disc diffusion | - | low |
| Amare et al.[10] | 2022 | Cross-sectional | Ethiopia | East Africa | Clinical | 290 | 68 | 17 | Disk diffusion | - | low |
| Bager et al.[11] | 2022 | - | Uganda | East Africa | Animal | 86 | 6 | 6 | Molecular | CTX-M-15, SHV-11 | Medium |
| Bayleyegn et al.[12] | 2021 | Cross-sectional | Ethiopia | East Africa | Clinical | 161 | 30 | 6 | Phenotypic | - | low |
| Bediako-Bowan et al.[13] | 2020 | - | Ghana | West Africa | Clinical | 4577 | 35 | 7 | Disk diffusion | - | low |
| Beshah et al.[14] | 2023 | Cross-sectional | Ethiopia | East Africa | Clinical | 1486 | 75 | 72 | Disk diffusion and E-test | - | low |
| Beyene et al.[15] | 2019 | Cross-sectional | Ethiopia | East Africa | Clinical | 947 | 72 | 55 | Disk diffusion | | low |
| Bitew et al.[16] | 2020 | Longitudinal | Ethiopia | East Africa | Clinical | 440 | 56 | 23 | Vitek 2 compact system | - | low |

| | | | | | | | | | | | |
|---|---|---|---|---|---|---|---|---|---|---|---|
| Buys et al.[17] | 2016 | Cross-sectional | South Africa | South Africa | Clinical | 410 | - | 339 | - | - | low |
| Chah et al.[18] | 2018 | Longitudinal | Nigeria | West Africa | Animal | 510 | 7 | 7 | Disk diffusion | SHV, CTX-M, TEM | low |
| Chinowaita et al.[19] | 2020 | - | Zimbabwe | South Africa | Clinical | 142 | 7 | 4 | Disk diffusion | - | low |
| Choonara et al.[20] | 2022 | Cross-sectional | Malawi | East Africa | Clinical | 694 | 29 | 20 | Phenotypic | - | low |
| Chukwu et al.[21] | 2022 | - | Nigeria | West Africa | Clinical | 499 | 15 | 13 | Phenotypic and genotypic assay | TEM and SHV | high |
| Damiano et al.[22] | 2021 | Cross-sectional | Tanzania | East Africa | Clinical | 35 | 20 | 17 | Disc combination | - | low |
| Darboe et al.[23] | 2023 | Cross-sectional | Gambia | West Africa | Clinical | 14,776 | 96 | 34 | Phenotypic | - | low |
| Dayie et al.[24] | 2022 | Cross-sectional | Ghana | West Africa | Clinical | 100 | 19 | 4 | Kirby-Bauer disc diffusion technique | - | low |
| Dela et al.[25] | 2022 | Cross-sectional | Ghana | West Africa | Clinical | 122 | 14 | 5 | Phenotypic and genotypic assay | TEM, CTX-M, SHV | high |

| | | | | | | | | | | | |
|---|---|---|---|---|---|---|---|---|---|---|---|
| Desta et al.[26] | 2016 | Cross-sectional | Ethiopia | East Africa | Clinical | 267 | 58 | 44 | Vitek 2 compact system | - | low |
| Dikoumba et al.[27] | 2021 | Longitudinal | Gabon | Central Africa | Clinical | 974 | - | 38 | agar disk diffusion and double-disk synergy test | SHV, CTX-M, TEM | low |
| Dirar et al.[28] | 2020 | - | Sudan | East Africa | Clinical | 171 | - | 58 | Phenotypic and genotypic assay | SHV, CTX-M, TEM | high |
| Djim-Adjim-ngana et al.[29] | 2020 | - | Cameroon | West Africa | Clinical | 57 | 5 | 4 | Disk diffusion | - | low |
| Dougnon et al.[30] | 2021 | Longitudinal | Benin | West Africa | Environment | 130 | 4 | 3 | disk diffusion and double disc diffusion | SHV | low |
| Eibach et al.[31] | 2016 | Cross-sectional | Ghana | West Africa | Clinical | 7172 | 41 | 34 | Phenotypic and genotypic assay | CTX-M | high |

| | | | | | | | | | | | |
|---|---|---|---|---|---|---|---|---|---|---|---|
| Eibach et al.[32] | 2018 | Cross-sectional | Ghana | West Africa | Animal | 200 | - | 35 | VITEK 2 system, combined disc test and molecular | CTX-M-15, CTX-M-1, CTX-M-2, CTX-M-14, SHV-12. | high |
| Endaylalu et al.[33] | 2020 | Cross-sectional | Ethiopia | East Africa | Clinical | 236 | 16 | 2 | Disk diffusion | - | low |
| Engda et al.[34] | 2018 | Cross-sectional | Ethiopia | East Africa | Environment | 384 | - | 24 | Disk diffusion | - | low |
| Erb et al.[35] | 2018 | - | Tanzania | East Africa | Environment | 636 | 7 | 1 | Phenotypic and genotypic assay | CTX-M-1 group | high |
| Fadare and Okoh[36] | 2021 | Cross-sectional | South Africa | South Africa | Environment | 6 | 9 | 5 | disk diffusion, conventional singleplex, duplex and multiplex PCR | TEM, OXA-1, OXA-48, CTX-M-1, CTX-M-2, SHV | high |
| Fadare et al.[37] | 2020 | Cross-sectional | South Africa | South Africa | Environment | 48 | 8 | 8 | Kirby-Bauer disk diffusion, Singleplex, duplex, | SHV, CTX-M2, OXA-48, CTX-M1, TEM, OXA-1 | high |

| | | | | | | | | | | | |
|---|---|---|---|---|---|---|---|---|---|---|---|
| | | | | | | | | | and multiplex PCR protocols | | |
| Fenta et al.[38] | 2020 | Cross-sectional | Ethiopia | East Africa | Clinical | 50 | 7 | 1 | Kirby-Bauer disc diffusion; double-disk diffusion method | - | low |
| Founou et al.[39] | 2019 | Cross-sectional | Cameroon | West Africa | Clinical | 53 | - | 10 | Phenotypic | - | low |
| Founou et al.[40] | 2018 | Cross-sectional | South Africa | South Africa | Clinical | 646 | - | 8 | Molecular | CTX-M-gp9, CTX-M-gp1, SHV, CTX-M-gp8/25, OXA-1-Like, TEM | medium |
| Founou et al.[41] | 2019 | Longitudinal | South Africa | South Africa | Clinical | 45 | - | 8 | Vitek® 2 System and Vitek® 2 Gram-negative Susceptibility card | SHV, CTX-M, TEM | high |

| | | | | | | | | | | | |
|---|---|---|---|---|---|---|---|---|---|---|---|
| | | | | | | | | | (AST-N255) and whole genome | | |
| Godonou et al.[42] | 2022 | Cross-sectional | Togo | West Africa | Clinical | 105 | - | 24 | Kirby-Bauer disk diffussion | - | low |
| Henson et al.[43] | 2017 | Longitudinal | Kenya | East Africa | Clinical | 221 | 221 | 198 | Double disk diffusion and whole genome sequencing | SHV and OKP | high |
| Ibrahim et al.[44] | 2017 | Cross-sectional | Nigeria | West Africa | Clinical | 248 | 108 | 68 | Double disc synergy test | - | low |
| Irenge et al.[45] | 2023 | Cross-sectional | DRC | | Clinical | 2051 | 449 | 89 | Phenotypic and genotypic assay | CTX-M (15), SHV, TEM | high |
| Irenge et al.[46] | 2014 | Cross-sectional | DRC | Central Africa | Clinical | 643 | 146 | 18 | Disk diffusion and Whole-genome sequencing | CTXM-1 | high |

| | | | | | | | | | | | |
|---|---|---|---|---|---|---|---|---|---|---|---|
| Isendahl et al.[47] | 2012 | Cross-sectional | Guinea-Bissau | West Africa | Clinical | 408 | 133 | 91 | Disk diffusion and Molecular | CTX-M-1, CTX-M-15 | high |
| Jesumirhewe et al.[48] | 2020 | Cross-sectional | Nigeria | West Africa | Clinical | 217 | - | 16 | Phenotypic and genotypic assay | CTX-M(15, 11), SHV(28) | high |
| Kaduma et al.[49] | 2019 | Longitudinal | Tanzania | East Africa | Clinical | 393 | 25 | 4 | Conventional disc diffusion | - | low |
| Kagia et al.[50] | 2019 | Cross-sectional | Kenya | East Africa | Clinical | 597 | - | 31 | Phenotypic | - | low |
| Kajeguka et al.[51] | 2015 | - | Tanzania | East Africa | Clinical | 330 | 55 | 22 | Disk diffusion | - | low |
| Kasew et al.[52] | 2021 | Cross-sectional | Ethiopia | East Africa | Clinical | 300 | 7 | 3 | Disk diffusion | - | low |
| Kateregga et al.[53] | 2015 | Cross-sectional | Uganda | East Africa | Clinical | 245 | 33 | 24 | Disk diffusion | - | low |
| Kenga et al.[54] | 2021 | Longitudinal | Mozambique | South Africa | Clinical | 97 | 3 | 2 | Kirby-Bauer disk diffussion | - | low |
| Kibwana et al.[55] | 2020 | Cross-sectional | Tanzania | East Africa | Clinical | 196 | - | 37 | Disk diffusion | - | low |
| Kibwana et al.[56] | 2023 | Cross-sectional | Tanzania | East Africa | Clinical | 142 | 35 | 3 | disk diffusion, | - | high |

| | | | | | | | | | | | |
|---|---|---|---|---|---|---|---|---|---|---|---|
| | | | | | | | | | E-test and molecular | | |
| Kibwana et al.[57] | 2022 | Cross-sectional | Tanzania | East Africa | Clinical | 200 | 46 | 46 | Disc diffusion and PCR | CTX-M and CTX-M-15 | high |
| Kiros et al.[58] | 2023 | Cross-sectional | Ethiopia | East Africa | Clinical | 383 | 51 | 34 | Phenotypic | - | low |
| Leski et al.[59] | 2016 | Cross-sectional | Sierra Leone | West Africa | Environment | 189 | 2 | 1 | Phenotypic and genotypic assay | CTX-M | high |
| Leski et al.[59] | 2016 | Cross-sectional | Sierra Leone | West Africa | Clinical | 93 | 15 | 9 | Phenotypic and genotypic assay | CTX-M | high |
| Letara et al.[60] | 2021 | Cross-sectional | Tanzania | East Africa | Clinical | 609 | - | 14 | Disk diffusion | - | low |
| Magale et al.[61] | 2015 | Cross-sectional | Kenya | East Africa | Clinical | 948 | 33 | 26 | - | - | low |
| Mahamat et al.[62] | 2019 | - | Chad | Central Africa | Clinical | 200 | 89 | 16 | Disk diffusion and Molecular | CTX-M-15 and CTX-M-14 | high |
| Mahamat et al.[63] | 2019 | - | Chad | Central Africa | Clinical | 1713 | - | 20 | Disk diffusion and Molecular | CTX-M-15, CTX-M-14, CTX-M-15/TEM-1, CTX- | high |

| | | | | | | | | | | | |
|---|---|---|---|---|---|---|---|---|---|---|---|
| | | | | | | | | | | M-15/OXA-1, CTX-M-15/TEM-1/OXA-1 | |
| Manyahi et al.[64] | 2014 | Decriptive cross-sectional | Tanzania | East Africa | Clinical | 100 | - | 11 | Disk diffusion | - | low |
| Manyahi et al.[65] | 2022 | Cross-sectional | Tanzania | East Africa | Clinical | 198 | 39 | 39 | Phenotypic | - | low |
| Mayanja et al.[66] | 2023 | Cross-sectional | Uganda | East Africa | Clinical | 273 | 50 | 25 | Disk diffusion and Molecular | CTXM-U/15, CTX-M, TEM, SHV | high |
| Mofolorunsho et al.[67] | 2021 | Cross-sectional | Nigeria | West Africa | Clinical | 200 | 28 | 24 | double disc | - | low |
| Moges et al.[68] | 2019 | - | Ethiopia | East Africa | Clinical | 532 | 97 | 79 | Disk diffusion | - | low |
| Moirongo et al.[69] | 2020 | - | Burkina Faso, Gabon, Ghana, and Tanzania | West Africa, central Africa, East Africa | Clinical | 3012 | 22 | 14 | Disk diffusion and Multilocus sequence typing | CTX-M15 | high |

| | | | | | | | | | | | |
|---|---|---|---|---|---|---|---|---|---|---|---|
| Montso et al.[70] | 2019 | Cross-sectional | South Africa | South Africa | Animal | 151 | 114 | 82 | disk-diffusion method; PCR | TEM, SHV, CTX-M, OXA | high |
| Moremi et al.[71] | 2017 | Cross-sectional | Tanzania | East Africa | Clinical | 107 | - | 6 | Disk diffusion and Molecular | SHV-1, SHV-11, CTX-M-15, CTX-M-9, TEM-1 | high |
| Motayo et al.[72] | 2013 | - | Nigeria | West Africa | Clinical | 97 | 30 | 5 | Disk diffusion | - | low |
| Moyo et al.[73] | 2020 | Cross-sectional | Tanzania | East Africa | Clinical | 2226 | 62 | 56 | Phenotypic | - | low |
| Mshana et al.[74] | 2016 | Cross-sectional | Tanzania | East Africa | Clinical | 334 | 53 | 2 | Disk diffusion and Molecular | CTX-M-15 | high |
| Mshana et al.[75] | 2013 | Cross-sectional | Tanzania | East Africa | Clinical | 1260 | 103 | 92 | Disc diffusion, double-disc synergy, PCR | CTX-M-15, TEM-1, SHV-11, TEM-104, TEM-176 | high |
| Mulinganya et al.[76] | 2021 | Cross-sectional | DRC | Central Africa | Clinical | 150 | 9 | 8 | Disk diffusion | - | low |
| Müller-Schulte et al.[77] | 2020 | Cross-sectional | Côte d’Ivoire | West Africa | Clinical | 107 | 107 | 90 | Disc diffusion, double- | - | low |

| | | | | | | | | | | | |
|---|---|---|---|---|---|---|---|---|---|---|---|
| | | | | | | | | | disc diffusion | | |
| Musicha et al.[78] | 2019 | Cross-sectional | Malawi | East Africa | Clinical | 205 | 205 | 34 | Disk diffusion and molecular | CTX-M-15, SHV-27, OXA-10, TEM-63, and SHV-7 | high |
| Mutua et al.[79] | 2023 | Cross-sectional | Kenya | East Africa | Clinical | 120 | - | 15 | Phenotypic and genotypic assay | CTX-M, OXA, SHV, TEM | high |
| Naas et al.[80] | 2016 | Cross-sectional | Madagascar | East Africa | Clinical | 808 | - | 15 | Phenotypic and genotypic assay | TEM, CTX-M-15 | high |
| Najjuka et al.[81] | 2016 | Cross-sectional | Uganda | East Africa | Clinical | 1448 | 10 | 10 | Disk diffusion and E-test | - | low |
| Nguekap et al.[82] | 2020 | prospective study | Cameroon | West Africa | Clinical | 110 | - | 4 | Disk diffusion | - | low |
| Nnaji et al.[83] | 2021 | Cross-sectional | Nigeria | West Africa | Animal | 450 | 8 | 8 | Disc diffusion | - | low |
| Obeng-Nkrumah et al.[84] | 2023 | Cross-sectional | Ghana | West Africa | Clinical | 382 | 9 | 2 | Kirby-Bauer disc diffusion, Double disk | SHV-31 | high |

| | | | | | | | | | | | |
|---|---|---|---|---|---|---|---|---|---|---|---|
| | | | | | | | | | diffusion tests and genotypic assay | | |
| Ogunbosi et al.[85] | 2020 | Cross-sectional | South Africa | South Africa | Clinical | 200 | 104 | 65 | Disk diffusion and Molecular | SHV, TEM | high |
| Olalekan et al.[86] | 2020 | Cross-sectional | Nigeria | West Africa | Clinical | 387 | 62 | 62 | Double-disk diffusion Test; whole-genome sequencing | CTX-M-14 | high |
| Olasehinde et al.[87] | 2021 | - | Nigeria | West Africa | Clinical | 1000 | - | 76 | disk diffusion method and molecular techniques | - | high |
| Ombelet et al.[88] | 2022 | Retrospective survailance | Benin | West Africa | Clinical | 383 | 58 | 36 | Disk diffusion and E-tests, double disk testing | - | low |

| | | | | | | | | | | | |
|---|---|---|---|---|---|---|---|---|---|---|---|
| Osei et al.[89] | 2022 | Cross-sectional | Ghana | West Africa | Clinical | 410 | 14 | 3 | Disk diffusion | - | low |
| Ouedraogo et al.[90] | 2016 | - | Burkina Faso, Gabon, Ghana, and Tanzania | West Africa | Clinical | 1602 | 70 | 46 | Disk diffusion and Molecular | CTX-M-15, CTX-M-27, and CTX-M-14 | high |
| Owusu et al.[91] | 2023 | Cross-sectional | Ghana | West Africa | Clinical | 181 | 30 | 7 | Phenotypic and genotypic assay | CTX-M, TEM, SHV | high |
| Pillay et al.[92] | 2021 | - | South Africa | South Africa | Clinical | 681 | 79 | 52 | Disk diffusion | - | low |
| Raji et al.[93] | 2013 | Cross-sectional | Nigeria | West Africa | Clinical | 102 | 32 | 12 | E-test | - | low |
| Rakotondrasoa et al.[94] | 2020 | - | Madagascar | East Africa | Clinical | 4001 | 251 | 65 | Disk diffusion, Molecular | CTX-M-15 | high |
| Rakotonirina et al.[95] | 2013 | Cross-sectional | Madagascar | East Africa | Clinical | 909 | 95 | 14 | Disc diffusion Method; PCR | CTX-M-15, SHV-12, TEM-1, and OXA-1 | high |
| Ramsamy et al.[96] | 2013 | Longitudinal | Republic of South Africa | South Africa | Clinical | 227 | 24 | 6 | standard laboratory techniques. | - | low |

| | | | | | | | | | | | |
|---|---|---|---|---|---|---|---|---|---|---|---|
| Sader et al.[97] | 2013 | Cross-sectional | South Africa | South Africa | Clinical | 2351 | 92 | 33 | broth microdilution | - | low |
| Sah et al.[98] | 2022 | Cross-sectional | Ghana | West Africa | Clinical | 258 | 71 | 27 | Disc diffusion Method; PCR | - | high |
| Sahle et al.[99] | 2022 | Cross-sectional | Ethiopia | East Africa | Clinical | 384 | 47 | 24 | Phenotypic | - | low |
| Sammarro et al.[100] | 2023 | Cross-sectional | Malawi | East Africa | Clinical | 2493 | 296 | 296 | CHROMagar isolation method; PCR | - | high |
| Sangare et al.[101] | 2016 | Cross-sectional | Mali | West Africa | Clinical | 2105 | 14 | 10 | Kirby-Bauer method, synergy test | - | low |
| Sanke-Waigana et al.[102] | 2021 | Retrospective study | Centra Africa Republic | Central Africa | Clinical | 7149 | - | 119 | Disk diffusion and Molecular | CTX-M (CTX-M1, CTX-M2, CTX-M8 CTX-M9), TEM, and SHV | high |

| | | | | | | | | | | | |
|---|---|---|---|---|---|---|---|---|---|---|---|
| Schaumburg et al.[103] | 2013 | Cross-sectional | Gabon | Central Africa | Clinical | 200 | - | 53 | Phenotypic and genotypic assay | CTX-M (1,8,15), TEM | high |
| Scherbaum et al.[104] | 2014 | Longitudinal | Gabon | Central Africa | Clinical | 187 | 6 | 3 | Micro dilution | - | low |
| Selamyhun et al.[105] | 2022 | Cross-sectional | Ethiopia | East Africa | Clinical | 384 | 31 | 18 | Disk diffusion | - | low |
| Seni et al.[106] | 2013 | Cross-sectional | Uganda | East Africa | Clinical | 216 | 39 | 36 | Standard disc diffusion technique | - | low |
| Sewunet et al.[107] | 2022 | Cross-sectional | Ethiopia | East Africa | Clinical | 1087 | 146 | 112 | Double-disk diffusion Test; whole-genome sequencing | - | high |
| Soré et al.[108] | 2021 | Cross-sectional | Burkina Faso | West Africa | Clinical | 149 | - | 18 | Disk diffusion | - | low |
| Soré et al..[109] | 2020 | - | Burkina Faso | West Africa | Environment | 293 | - | 42 | Disk diffusion | - | low |
| Teferi et al.[110] | 2023 | Cross-sectional | Ethiopia | East Africa | Clinical | 386 | 21 | 6 | Kirby-Bauer disc diffusion | - | low |

| | | | | | | | | | | | |
|---|---|---|---|---|---|---|---|---|---|---|---|
| Teklu et al.[111] | 2019 | Cross-sectional | Ethiopia | East Africa | Clinical | 426 | 108 | 81 | Disk diffusion | - | low |
| Tellevik et al.[112] | 2016 | - | Tanzania | East Africa | Clinical | 603 | - | 139 | Disk diffusion and Molecular | CTX-M-15-like | high |
| Tesfaye et al.[113] | 2019 | Cross-sectional | Ethiopia | East Africa | Environment | 24 | 8 | 1 | Phenotypic | - | low |
| Tola et al.[114] | 2021 | Cross-sectional | Ethiopia | East Africa | Clinical | 269 | 39 | 8 | VITEK 2 ESBL test | - | low |
| Tufa et al.[115] | 2022 | Prospective | Ethiopia | East Africa | Clinical | 761 | 6 | 6 | Phenotypic and genotypic assay | SHV, CTX-M, TEM | high |
| Tumuhamye et al.[116] | 2021 | Cross-sectional | Uganda | East Africa | Clinical | 1472 | 145 | 16 | Phenotypic | - | low |
| Yehouenou et al.[117] | 2020 | Cross-sectional | Benin | West Africa | Clinical | 304 | 30 | 25 | Phenotypic | - | low |
| Zemtsa et al.[118] | 2022 | Cross-sectional | Cameroon | West Africa | Clinical | 120 | 7 | 4 | Phenotypic and genotypic assay | CTX-M (1,2,9), TEM, | high |

Country-specific prevalence

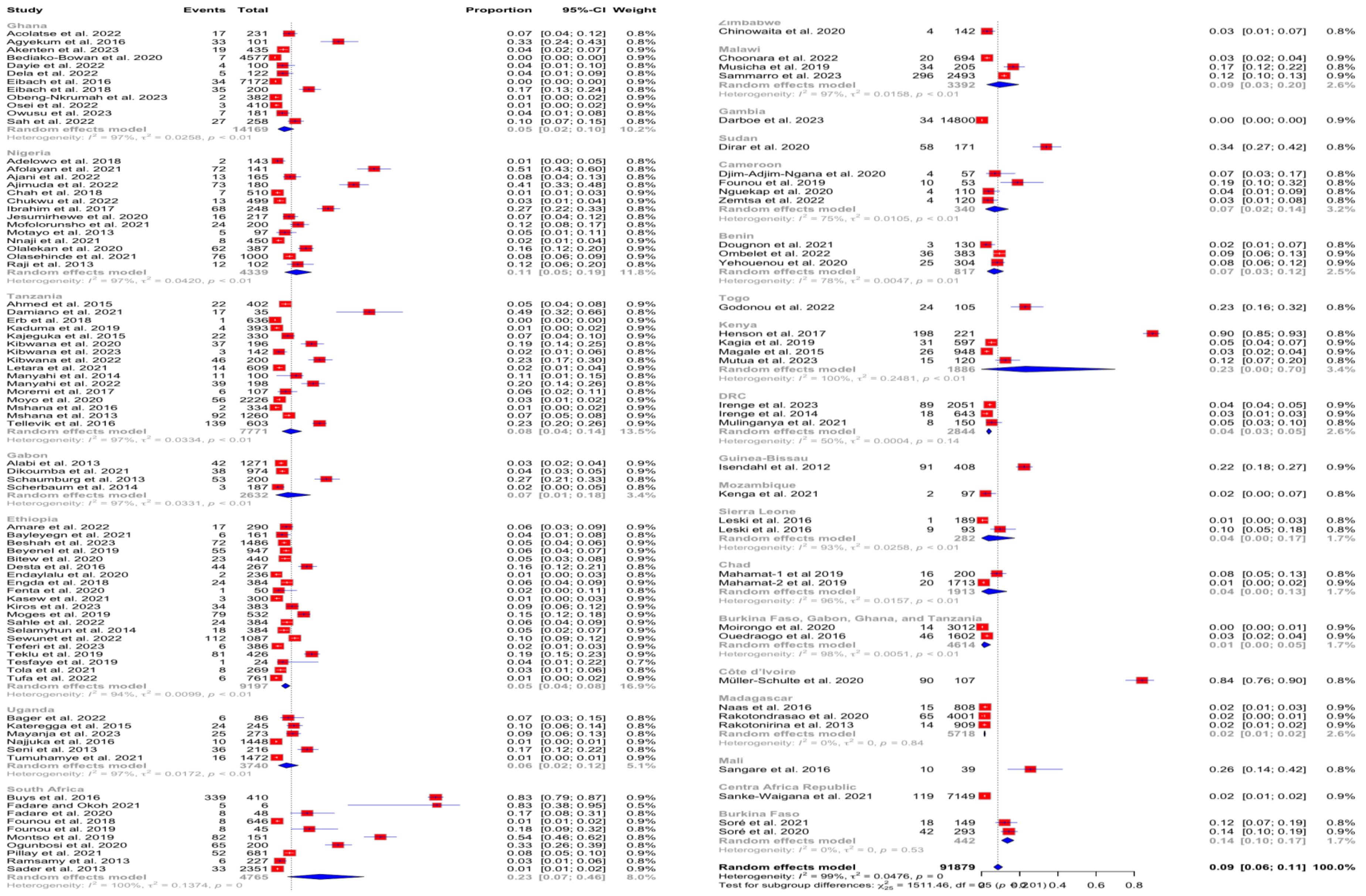


**Supplementary Figure 2**. Forest plot showing subgroup analysis of ESBL-producing *K. pneumoniae* based on countries in sub-Saharan Africa

FUNNEL PLOT – FOR PUBLICATION BIAS

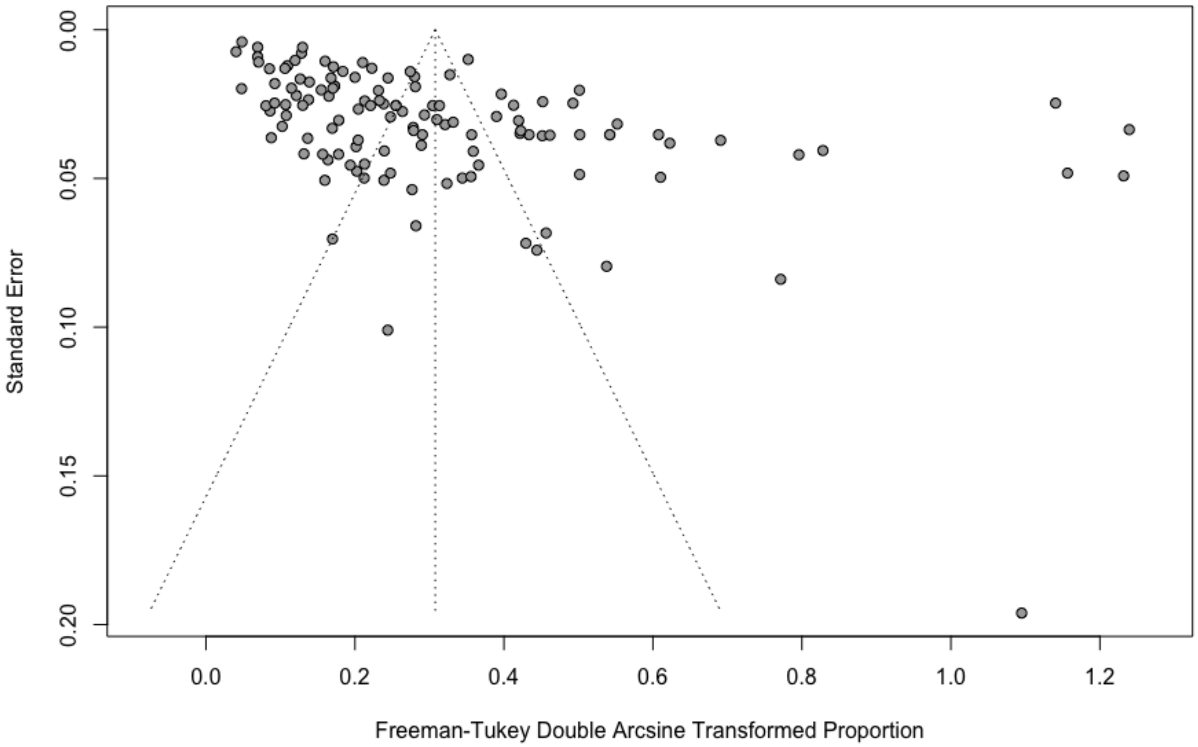


**Supplementary Figure 3.** Funnel plot of ESBL-producing *K. pneumoniae* studies in sub-Saharan Africa

**Supplementary Material References of eligible articles included in this study (Table 1)**


1 Acolatse JEE, Portal EAR, Boostrom I, Akafity G, Dakroah MP, Chalker VJ, et al. Environmental surveillance of ESBL and carbapenemase-producing gram-negative bacteria in a Ghanaian Tertiary Hospital. *Antimicrob Resist Infect Control* 2022; **11**: 1-15

2 Adelowo OO, Caucci S, Banjo OA, Nnanna OC, Awotipe EO, Peters FB, et al. Extended Spectrum Beta-Lactamase (ESBL)-producing bacteria isolated from hospital wastewaters, rivers and aquaculture sources in Nigeria. *Environmental Science and Pollution Research* 2018; **25**: 2744–55.

3 Afolayan AO, Oaikhena AO, Aboderin AO, et al. Clones and Clusters of Antimicrobial-Resistant Klebsiella from Southwestern Nigeria. *Clinical Infectious Diseases* 2021; **73**: S308–15.

4 Agyekum A, Fajardo-Lubián A, Ansong D, Partridge SR, Agbenyega T, Iredell JR. blaCTX-M-15 carried by IncF-type plasmids is the dominant ESBL gene in Escherichia coli and *Klebsiella pneumoniae* at a hospital in Ghana. *Diagn Microbiol Infect Dis* 2016; **84**: 328–33.

5 Ahmed M, Moremi N, Mirambo MM, et al. Multi-resistant gram negative enteric bacteria causing urinary tract infection among malnourished underfives admitted at a tertiary hospital, northwestern, Tanzania. *Ital J Pediatr* 2015; **19**:41.

6 Ajani TA, Elikwu CJ, Anaedobe CG et al. Evaluation of phenotypic and molecular technique in the detection of extended spectrum beta-lactamase (esbl)-producing gram negative bacilli in Ogun state, Nigeria. *Ann Ibd Pg Med* 2022; **20**: 160-68.

7 Ajimuda OE, Sanmi-Kayode I, Adeniyi OO, Alaka OO, Onipede A. Prevalence of extended spectrum Beta-Lactamase producing Klebsiella species from patients' specimens in a tertiary teaching hospital in Ile-Ife, Southwest Nigeria. *Afr Health Sci* 2022; **22**: 146–55.

8 Akenten CW, Khan NA, Mbwana J, et al. Carriage of ESBL-producing *Klebsiella pneumoniae* and Escherichia coli among children in rural Ghana: a cross-sectional study. *Antimicrob Resist Infect Control* 2023; **12**: 60

9 Alabi AS, Frielinghaus L, Kaba H, et al. Retrospective analysis of antimicrobial resistance and bacterial spectrum of infection in Gabon, Central Africa. *BMC Infect Dis* 2013; **13**: 455

10 Amare A, Eshetie S, Kasew D, Moges F. High prevalence of fecal carriage of Extended-spectrum beta-lactamase and carbapenemase-producing Enterobacteriaceae among food handlers at the University of Gondar, Northwest Ethiopia. *PLoS One* 2022; **17**: e0264818

11 Bager SL, Kakaala I, Kudirkiene E, Byarugaba DK, Olsen JE. Genomic characterization of multidrug-resistant extended-spectrum β-lactamase–producing *Escherichia coli* and *Klebsiella pneumoniae* from chimpanzees (pan troglodytes) from wild and sanctuary locations in Uganda. *J Wildl Dis* 2022; **58**: 269–78.

12 Bayleyegn B, Fisaha R, Kasew D. Fecal carriage of extended spectrum beta-lactamase producing Enterobacteriaceae among HIV infected children at the University of Gondar Comprehensive *Specialized* Hospital Gondar, Ethiopia. *AIDS Res Ther* 2021; **18**: 19

13 Bediako-Bowan AAA, Kurtzhals JAL, Mølbak K, Labi AK, Owusu E, Newman MJ. High rates of multi-drug resistant gram-negative organisms associated with surgical site infections in a teaching hospital in Ghana. *BMC Infect Dis* 2020; **20**: 890

14 Beshah D, Desta AF, Woldemichael GB, et al. High burden of ESBL and carbapenemase-producing gram-negative bacteria in bloodstream infection patients at a tertiary care hospital in Addis Ababa, Ethiopia. *PLoS One* 2023; **18**: e0287453

15 Beyene D, Bitew A, Fantew S, Mihret A, Evans M. Multidrug-resistant profile and prevalence of extended spectrum β-lactamase and carbapenemase production in fermentative Gram-negative bacilli recovered from patients and specimens referred to National Reference Laboratory, Addis Ababa, Ethiopia. *PLoS One* 2019; **14**: e0222911

16 Bitew A, Tsige E. High Prevalence of Multidrug-Resistant and Extended-Spectrum β -Lactamase-Producing Enterobacteriaceae: A Cross-Sectional Study at Arsho Advanced Medical Laboratory, Addis Ababa, Ethiopia. *J Trop Med* 2020; **2020**: 6167234

17 Buys H, Muloiwa R, Bamford C, Eley B. *Klebsiella pneumoniae* bloodstream infections at a South African children's hospital 2006-2011, a cross-sectional study. *BMC Infect Dis* 2016; **16**: 570

18 Chah KF, Ugwu IC, Okpala A, et al. Detection and molecular characterisation of extended-spectrum β-lactamase-producing enteric bacteria from pigs and chickens in Nsukka, Nigeria. *J Glob Antimicrob Resist* 2018; **15**: 36–40.

19 Chinowaita F, Chaka W, Nyazika TK, et al. Sepsis in cancer patients residing in Zimbabwe: Spectrum of bacterial and fungal aetiologies and their antimicrobial susceptibility patterns. *BMC Infect Dis* 2020; **20**: 161

20 Choonara FE, Haldorsen BC, Ndhlovu I, et al. Antimicrobial susceptibility profiles of clinically important bacterial pathogens at the Kamuzu Central Hospital in Lilongwe, Malawi. *Malawi Medical Journal* 2022; **34**: 9–16.

21 Chukwu EE, Awoderu OB, Enwuru CA, et al. High prevalence of resistance to third-generation cephalosporins detected among clinical isolates from sentinel healthcare facilities in Lagos, Nigeria. *Antimicrob Resist Infect Control* 2022; **11**: 134

22 Damiano P, Salema EJ, Silago V. The susceptibility of multidrug resistant and biofilm forming *Klebsiella pneumoniae* and escherichia coli to antiseptic agents used for preoperative skin preparations at zonal referral hospital in mwanza, Tanzania. *Malawi Medical Journal* 2021; **33**: 59–64.

23 Darboe S, Mirasol R, Adejuyigbe B, et al. Using an Antibiogram Profile to Improve Infection Control and Rational Antimicrobial Therapy in an Urban Hospital in The Gambia, Strategies and Lessons for Low- and Middle-Income Countries. *Antibiotics* 2023; **12**: 790

24 Dayie NT, Bannah V, Dwomoh FP, Kotey FC, Donkor ES. Distribution and Antimicrobial Resistance Profiles of Bacterial Aetiologies of Childhood Otitis Media in Accra, Ghana. *Microbiol Insights* 2022; **15**: 117863612211044.

25 Dela H, Egyir B, Majekodunmi AO, et al. Diarrhoeagenic *E. coli* occurrence and antimicrobial resistance of Extended Spectrum Beta-Lactamases isolated from diarrhoea patients attending health facilities in Accra, Ghana. *PLoS One* 2022; **17**: 0268991

26 Desta K, Woldeamanuel Y, Azazh A, et al. High gastrointestinal colonization rate with extended-spectrum β-lactamase-producing Enterobacteriaceae in hospitalized patients: Emergence of carbapenemase-producing *K. Pneumoniae* in Ethiopia. *PLoS One* 2016; **11**: e0161685

27 Dikoumba AC, Onanga R, Boundenga L, Bignoumba M, Ngoungou EB, Godreuil S. Prevalence and Characterization of Extended-Spectrum Beta-Lactamase-Producing Enterobacteriaceae in Major Hospitals in Gabon. *Microbial Drug Resistance* 2021; **27**: 1525–34.

28 Dirar MH, Bilal NE, Ibrahim ME, Hamid ME. Prevalence of extended-spectrum β-lactamase (Esbl) and molecular detection of blatem, blashv and blactx-m genotypes among enterobacteriaceae isolates from patients in khartoum, sudan. *Pan African Medical Journal* 2020; **37**: 1–11.

29 Djim-Adjim-ngana K, Oumar LA, Mbiakop BW, Njifon HLM, Crucitti T, Nchiwan EN, et al. Prevalence of extended-spectrum beta-lactamaseproducing enterobacterial urinary infections

and associated risk factors in small children of garoua, Northern Cameroon. *Pan African Medical Journal* 2020; **36**: 1–10.

30 Dougnon V, Houssou VMC, Anago E, Nanoukon C, Mohammed J, Agbankpe J, et al. Assessment of the Presence of Resistance Genes Detected from the Environment and Selected Food Products in Benin. *J Environ Public Health* 2021; **2021**: 8420590

31 Eibach D, Campos CB, Krumkamp R, et al. Extended spectrum beta-lactamase producing Enterobacteriaceae causing bloodstream infections in rural Ghana, 2007-2012. *International Journal of Medical Microbiology* 2016; **306**: 249–54.

32 Eibach D, Dekker D, Gyau Boahen K, et al. Extended-spectrum beta-lactamase-producing Escherichia coli and *Klebsiella pneumoniae* in local and imported poultry meat in Ghana. *Vet Microbiol* 2018; **217**: 7–12.

33 Endaylalu K, Abera B, Mulu W. Extended spectrum beta lactamase producing bacteria among outpatients with ear infection at FelegeHiwot Referral Hospital, North West Ethiopia. *PLoS One*. 2020; **15**: 0238891

34 Engda T, Moges F, Gelaw A, Eshete S, Mekonnen F. Prevalence and antimicrobial susceptibility patterns of extended spectrum beta-lactamase producing Entrobacteriaceae in the University of Gondar Referral Hospital environments, northwest Ethiopia. *BMC Res Notes* 2018; **11**: 335

35 Erb S, D'Mello-Guyett L, Malebo HM, et al. High prevalence of ESBL-Producing E. coli in private and shared latrines in an informal urban settlement in Dar es Salaam, Tanzania. *Antimicrob Resist Infect Control* 2018; **7**: 3

36 Fadare FT, Okoh AI. Distribution and molecular characterization of ESBL, pAmpC β-lactamases, and non-β-lactam encoding genes in Enterobacteriaceae isolated from hospital wastewater in Eastern Cape Province, South Africa. *PLoS One* 2021; **16**: e0254753

37 Fadare FT, Adefisoye MA, Okoh AI. Occurrence, identification, and antibiogram signatures of selected Enterobacteriaceae from Tsomo and Tyhume rivers in the Eastern Cape Province, Republic of South Africa. *PLoS One* 2020; **15**: 0238084

38 Fenta A, Dagnew M, Eshetie S, Belachew T. Bacterial profile, antibiotic susceptibility pattern and associated risk factors of urinary tract infection among clinically suspected children attending at Felege-Hiwot comprehensive and specialized hospital, Northwest Ethiopia. A prospective study. *BMC Infect Dis* 2020; **20**: 673

39 Founou LL, Founou RC, Ntshobeni N, et al. Emergence and spread of extended spectrum β-lactamase producing enterobacteriaceae (ESBL-PE) in pigs and exposed workers: A multicentre comparative study between Cameroon and South Africa. *Pathogens* 2019; **8**: 10

40 Founou RC, Founou LL, Essack SY. Extended spectrum beta-lactamase mediated resistance in carriage and clinical gram-negative ESKAPE bacteria: A comparative study between a district and tertiary hospital in South Africa. *Antimicrob Resist Infect Control* 2018; **7**: 134

41 Founou RC, Founou LL, Allam M, Ismail A, Essack SY. Whole Genome Sequencing of Extended Spectrum β-lactamase (ESBL)-producing *Klebsiella pneumoniae* Isolated from Hospitalized Patients in KwaZulu-Natal, South Africa. *Sci Rep* 2019; **9**: 6266

42 Godonou AM, Lack F, Gbeasor-Komlanvi FA, et al. High faecal carriage of extended-spectrum beta-lactamase producing Enterobacteriaceae (ESBL-PE) among hospitalized patients at Sylvanus Olympio Teaching Hospital, Lomé, Togo in 2019. *African Journal of Clinical and Experimental Microbiology* 2022; **23**: 40–8.

43 Henson SP, Boinett CJ, Ellington MJ, et al. Molecular epidemiology of *Klebsiella pneumoniae* invasive infections over a decade at Kilifi County Hospital in Kenya. *International Journal of Medical Microbiology* 2017; **307**:422–9.

44 Ibrahim Y, Sani Y, Saleh Q, Saleh A, Hakeem G. Phenotypic Detection of Extended Spectrum Beta lactamase and Carbapenemase Co-producing Clinical Isolates from Two Tertiary Hospitals in Kano, North West Nigeria. *Ethiop J Health Sci* 2017; **27**: 3–10.

45 Irenge LM, Ambroise J, Bearzatto B, Durant JF, Bonjean M, Gala JL. Genomic Characterization of Multidrug-Resistant Extended Spectrum β-Lactamase-Producing *Klebsiella pneumoniae* from Clinical Samples of a Tertiary Hospital in South Kivu Province, Eastern Democratic Republic of Congo. *Microorganisms* 2023; **11**: 525

46 Irenge LM, Kabego L, Vandenberg O, Chirimwami RB, Gala JL. Antimicrobial resistance in urinary isolates from inpatients and outpatients at a tertiary care hospital in South-Kivu Province (Democratic Republic of Congo). *BMC Res Notes* 2014; **7**: 374

47 Isendahl J, Turlej-Rogacka A, Manjuba C, Rodrigues A, Giske CG, Nauclér P. Fecal Carriage of ESBL-Producing E. coli and K. pneumoniae in Children in Guinea-Bissau: A Hospital-Based Cross-Sectional Study. *PLoS One* 2012; **7**: e51981

48 Jesumirhewe C, Springer B, Allerberger F, Ruppitsch W. Whole genome sequencing of extendedspectrum β-lactamase genes in Enterobacteriaceae isolates from Nigeria. *PLoS One* 2020; **15**: 0231146

49 Kaduma J, Seni J, Chuma C, et al. Urinary tract infections and preeclampsia among pregnant women attending two hospitals in Mwanza City, Tanzania: A 1:2 Matched case-control study. *Biomed Res Int* 2019; **2019**: 3937812

50 Kagia N, Kosgei P, Ooko M, et al. Carriage and Acquisition of Extended-spectrum β-Lactamase-producing Enterobacterales among Neonates Admitted to Hospital in Kilifi, Kenya. *Clinical Infectious Diseases* 2019; **69**: 751–9.

51 Kajeguka DC, Nambunga PP, Kabissi F, et al. Antimicrobial resistance patterns of phenotype Extended Spectrum Beta-Lactamase producing bacterial isolates in a referral hospital in northern Tanzania. *Tanzan J Health Res* 2015; **17**: 10.4314/thrb.v17i3.2

52 Kasew D, Eshetie S, Diress A, Tegegne Z, Moges F. Multiple drug resistance bacterial isolates and associated factors among urinary stone patients at the University of Gondar Comprehensive Specialized Hospital, Northwest Ethiopia. *BMC Urol* 2021; **21**: 27

53 Kateregga JN, Kantume R, Atuhaire C, Lubowa MN, Ndukui JG. Phenotypic expression and prevalence of ESBL-producing Enterobacteriaceae in samples collected from patients in various wards of Mulago Hospital, Uganda. *BMC Pharmacol Toxicol* 2015; **16**: 14

54 Kenga DB, Gebretsadik T, Simbine S, et al. Community-acquired bacteremia among HIV-infected and HIV-exposed uninfected children hospitalized with fever in Mozambique. *International Journal of Infectious Diseases* 2021; **109**: 99–107.

55 Kibwana UO, Majigo M, Kamori D, Manyahi J. High fecal carriage of extended Beta Lactamase producing Enterobacteriaceae among adult patients admitted in referral hospitals in Dar es Salaam, Tanzania. *BMC Infect Dis* 2020; **20**: 557

56 Kibwana UO, Manyahi J, Sandnes HH, et al. Fluoroquinolone resistance among fecal extended spectrum βeta lactamases positive Enterobacterales isolates from children in Dar es Salaam, Tanzania. *BMC Infect Dis* 2023; **23**: 135

57 Kibwana UO, Manyahi J, Sandnes HH, et al. Gastrointestinal colonization of extended-spectrum beta-lactamase-producing bacteria among children below five years of age hospitalized with fever in Dar es Salaam, Tanzania. *J Glob Antimicrob Resist* 2022; **30**: 107–14.

58 Kiros T, Belete D, Andualem T, et al. Carriage of β-lactamase and carbapenemase-producing Enterobacteriaceae in hospitalized patients at debre tabor comprehensive specialized hospital. *Heliyon* 2023; **9**: e20072

59 Leski TA, Taitt CR, Bangura U, et al. High prevalence of multidrug resistant Enterobacteriaceae isolated from outpatient urine samples but not the hospital environment in Bo, Sierra Leone. *BMC Infect Dis* **2016**; **16**: 167

60 Letara N, Ngocho JS, Karami N, et al. Prevalence and patient related factors associated with Extended-Spectrum Beta-Lactamase producing Escherichia coli and *Klebsiella pneumoniae* carriage and infection among pediatric patients in Tanzania. *Sci Rep* 2021; **11**: 22759.

61 Magale HI, Kassim IA, Odera SA, Omolo MJ, Jaoko WG, Jolly PE. Antibiotic susceptibility of organisms causing urinary tract infection in patients presenting at Kenyatta national hospital, Nairobi. *East Afr Med J* 2015; **92**: 333-37.

62 Mahamat OO, Lounnas M, Hide M, et al. High prevalence and characterization of extended-spectrum ß-lactamase producing Enterobacteriaceae in Chadian hospitals. *BMC Infect Dis* 2019; **19**: 205

63 Mahamat OO, Tidjani A, Lounnas M, et al. Fecal carriage of extended-spectrum β-lactamase-producing Enterobacteriaceae in hospital and community settings in Chad. *Antimicrob Resist Infect Control* 2019; **8**: 169

64 Manyahi J, Matee MI, Majigo M, Moyo S, Mshana SE, Lyamuya EF. Predominance of multi-drug resistant bacterial pathogens causing surgical site infections in Muhimbili national hospital, Tanzania. *BMC Res Notes* 2014; **7**: 10.1186/1756-0500-7-500

65 Manyahi J, Majigo M, Kibwana U, Kamori D, Lyamuya EF. Colonization of Extended-spectrum β-lactamase producing Enterobacterales and meticillin-resistant S. aureus in the intensive care unit at a tertiary hospital in Tanzania: Implications for Infection control and prevention. *Infection Prevention in Practice* 2022; **4**: 100212.

66 Mayanja R, Muwonge A, Aruhomukama D, et al. Source-tracking ESBL-producing bacteria at the maternity ward of Mulago hospital, Uganda. *PLoS One* 2023; **18**: e0286955.

67 Mofolorunsho KC, Ocheni HO, Aminu RF, Omatola CA, Olowonibi OO. Prevalence and antimicrobial susceptibility of extended-spectrum beta lactamases-producing escherichia coli and *Klebsiella pneumoniae* isolated in selected hospitals of anyigba, *Nigeria. Afr Health Sci* 2021; **21**: 505–12.

68 Moges F, Eshetie S, Abebe W, et al. High prevalence of extended-spectrum beta-lactamase-producing Gram-negative pathogens from patients attending Felege Hiwot Comprehensive Specialized Hospital, Bahir Dar, Amhara region. *PLoS One* 2019; **14**: e0215177

69 Moirongo RM, Lorenz E, Ntinginya NE, et al. Regional Variation of Extended-Spectrum Beta-Lactamase (ESBL)-Producing Enterobacterales, Fluoroquinolone-Resistant Salmonella enterica and Methicillin-Resistant Staphylococcus aureus Among Febrile Patients in Sub-Saharan Africa. *Front Microbiol* 2020; **11**: 567235

70 Montso KP, Dlamini SB, Kumar A, Ateba CN, Garcia-Perdomo HA. Antimicrobial Resistance Factors of Extended-Spectrum Beta-Lactamases Producing Escherichia coli and *Klebsiella pneumoniae* Isolated from Cattle Farms and Raw Beef in North-West Province, South Africa. *Biomed Res Int* 2019; **2019**: 4318306

71 Moremi N, Claus H, Vogel U, Mshana SE. Faecal carriage of CTX-M extended-spectrum beta-lactamase-producing Enterobacteriaceae among street children dwelling in Mwanza city, Tanzania. *PLoS One* 2017; **12**: 0184592

72 Motayo BO, Akinduti PA, Adeyakinu FA, et al. Antibiogram and plasmid profiling of carbapenemase and extended spectrum beta-lactamase (ESBL) producing Escherichia coli and *Klebsiella pneumoniae* in Abeokuta, South Western, Nigeria. *Afr Health Sci* 2013; **13**: 1091–97.

73 Moyo SJ, Manyahi J, Blomberg B, et al. Bacteraemia, Malaria, and Case Fatality Among Children Hospitalized With Fever in Dar es Salaam, Tanzania. *Front Microbiol* 2020; **11**: 10.3389/fmicb.2020.02118

74 Mshana SE, Falgenhauer L, Mirambo MM, et al. Predictors of blaCTX-M-15 in varieties of Escherichia coli genotypes from humans in community settings in Mwanza, Tanzania. *BMC Infect Dis* 2016; **16**: 187

75 Mshana SE, Hain T, Domann E, Lyamuya EF, Chakraborty T, Imirzalioglu C. Predominance of *Klebsiella pneumoniae* ST14 carrying CTX-M-15 causing neonatal sepsis in Tanzania. *BMC Infect Dis* 2013; **13**: 466

76 Mulinganya GM, Claeys M, Balolebwami SZ, et al. Etiology of Early-Onset Neonatal Sepsis and Antibiotic Resistance in Bukavu, Democratic Republic of the Congo. *Clinical Infectious Diseases* 2021; **73**: E976–80.

77 Müller-Schulte E, Tuo MN, Akoua-Koffi C, Schaumburg F, Becker SL. High prevalence of ESBL-producing *Klebsiella pneumoniae* in clinical samples from central Côte d'Ivoire. *International Journal of Infectious Diseases* 2020; **91**: 207–9.

78 Musicha P, Msefula CL, Mather AE, et al. Genomic analysis of *Klebsiella pneumoniae* isolates from Malawi reveals acquisition of multiple ESBL determinants across diverse lineages. Journal of Antimicrobial Chemotherapy. 2019; **74**: 1223–32.

79 Mutua JM, Njeru JM, Musyoki AM. Extended-spectrum β-lactamase- producing gram-negative bacterial infections in severely ill COVID-19 patients admitted in a national referral hospital, Kenya. *Ann Clin Microbiol Antimicrob* 2023; **22**: 10.1186/s12941-023-00641-8

80 Naas T, Cuzon G, Robinson AL, et al. Neonatal infections with multidrug-resistant ESBL-producing E. cloacae and K. pneumoniae in Neonatal Units of two different Hospitals in Antananarivo, Madagascar. *BMC Infect Dis* 2016; **16**: 275

81 Najjuka CF, Kateete DP, Kajumbula HM, Joloba ML, Essack SY. Antimicrobial susceptibility profiles of Escherichia coli and *Klebsiella pneumoniae* isolated from outpatients in urban and rural districts of Uganda. *BMC Res Notes* 2016; **9**: 27113038

82 Nguekap WLN, Lontsi TI, Betbeui AC, Fewou SN. Carriage of extended-spectrum beta-lactamase-producing Enterobacteriaceae by healthy school children from two remote villages in western Cameroon. *Int J Biol Chem Sci* 2020; **14**: 3018–30.

83 Nnaji JO, Moses IB, Ejikeugwu PC, Nwakaeze EA, Ude-Ude I, Iroha IR. Antibiogram and molecular characterization of AmpC and ESBL-producing gram-negative bacteria from poultry and abattoir samples. *Pakistan Journal of Biological Sciences* 2021; **24**: 193–8.

84 Obeng-Nkrumah N, Hansen DS, Awuah-Mensah G, et al. High level of colonization with third-generation cephalosporin-resistant Enterobacterales in African community settings, Ghana. *Diagnostic Microbiology and Infectious Disease* 2023; **106**: 115918

85 Ogunbosi BO, Moodley C, Naicker P, Nuttall J, Bamford C, Eley B. Colonisation with extended spectrum betalactamase- producing and carbapenemresistant Enterobacterales in children admitted to a paediatric referral hospital in South Africa. *PLoS One* 2020; **15**: e0241776

86 Olalekan A, Onwugamba F, Iwalokun B, Mellmann A, Becker K, Schaumburg F. High proportion of carbapenemase-producing Escherichia coli and *Klebsiella pneumoniae* among extended-spectrum β-lactamase-producers in Nigerian hospitals. *J Glob Antimicrob Resist* 2020; **21**: 8–12.

87 Olasehinde O, Lamikanra A. Pattern of Esbls in Uro-Pathogens Obtained from a Nigerian Tertiary Hospital. *Nigerian Journal of Pharmaceutical Research* 2021; **16**: 139–47.

88 Ombelet S, Kpossou G, Kotchare C, et al. Blood culture surveillance in a secondary care hospital in Benin: epidemiology of bloodstream infection pathogens and antimicrobial resistance. *BMC Infect Dis* 2022; **22**: 119

89 Osei MM, Dayie NTKD, Azaglo GSK, et al. Alarming Levels of Multidrug Resistance in Aerobic Gram-Negative Bacilli Isolated from the Nasopharynx of Healthy Under-Five Children in Accra, Ghana. *Int J Environ Res Public Health* 2022; **19**: 10927

90 Ouedraogo AS, Sanou M, Kissou A, et al. High prevalence of extended-spectrum ß-lactamase producing enterobacteriaceae among clinical isolates in Burkina Faso. *BMC Infect Dis* 2016; **16**: 326

91 Owusu FA, Obeng-Nkrumah N, Gyinae E, et al. Occurrence of Carbapenemases, Extended-Spectrum Beta-Lactamases and AmpCs among Beta-Lactamase-Producing Gram-Negative Bacteria from Clinical Sources in Accra, Ghana. *Antibiotics* 2023; **12**: 1016

92 Pillay D, Naidoo L, Swe Swe-Han K, Mahabeer Y. Neonatal sepsis in a tertiary unit in South Africa. *BMC Infect Dis* 2021; **21**: 225

93 Raji MA, Jamal W, Ojemhen O, Rotimi VO. Point-surveillance of antibiotic resistance in Enterobacteriaceae isolates from patients in a Lagos Teaching Hospital, Nigeria. *J Infect Public Health* 2013; **6**: 431–7.

94 Rakotondrasoa A, Passet V, Herindrainy P, et al. Characterization of *Klebsiella pneumoniae* isolates from a mother-child cohort in Madagascar. *Journal of Antimicrobial Chemotherapy* 2020; **75**: 1736–46.

95 Rakotonirina HC, Garin B, Randrianirina F, Richard V, Talarmin A, Arlet G. Molecular characterization of multidrug-resistant extended-spectrum β-lactamase-producing Enterobacteriaceae isolated in Antananarivo, Madagascar. 2013; **13**: 1-10

96 Ramsamy Y, Muckart DJJ, Han KSS. Microbiological surveillance and antimicrobial stewardship minimise the need for ultrabroad-spectrum combination therapy for treatment of nosocomial infections in a trauma intensive care unit: An audit of an evidence-based empiric antimicrobial policy. *South African Medical Journal* 2013; **103**: 371–6.

97 Sader HS, Flamm RK, Jones RN. Antimicrobial activity of ceftaroline and comparator agents tested against bacterial isolates causing skin and soft tissue infections and community-acquired

respiratory tract infections isolated from the Asia-Pacific region and South Africa (2010). *Diagn Microbiol Infect Dis* 2013; **76**: 61–8.

98 Sah AK, Feglo PK. Plasmid-mediated quinolone resistance determinants in clinical bacterial pathogens isolated from the Western Region of Ghana: a cross-sectional study. *Pan Afr Med J* 2022; **43**: 207.

99 Sahle Z, Engidaye G, Shenkute D, Metaferia Y, Shibabaw A. High Prevalence of Multi-Drug Resistance and Extended-Spectrum Beta-Lactamase-Producing Enterobacteriaceae Among Hospitalized Patients Presumptive for Bacterial Infection at Debre Berhan Comprehensive Specialized Hospital, Ethiopia. *Infect Drug Resist* 2022; **15**: 2639–56.

100 Sammarro M, Rowlingson B, Cocker D, et al. Risk Factors, Temporal Dependence, and Seasonality of Human Extended-Spectrum β-Lactamases-Producing Escherichia coli and *Klebsiella pneumoniae* Colonization in Malawi: A Longitudinal Model-Based Approach. *Clinical Infectious Diseases*. 2023; **77**: 1-8

101 Sangare SA, Maiga AI, Guindo I, et al. Prevalence of ESBL-producing enterobacteriaceae isolated from blood cultures in Mali. *J Infect Dev Ctries* 2016; **10**: 1059–64.

102 Sanke-Waïgana H, Mbecko JR, Ngaya G, Manirakiza A, Alain BA. 2003-2019: Explosive spread of enterobacteria producing extended-spectrum beta-lactamases in Bangui Central African Republic. *Pan African Medical Journal* 2021; **39**: 22

103 Schaumburg F, Alabi A, Kokou C, et al. High burden of extended-spectrum β-lactamase-producing enterobacteriaceae in Gabon. *Journal of Antimicrobial Chemotherapy.* 2013; **68**: 2140–3.

104 Scherbaum M, Kösters K, Mürbeth RE, et al. Incidence, pathogens and resistance patterns of nosocomial infections at a rural hospital in Gabon. *BMC Infect Dis* 2014; **14**: 124

105 Selamyhun T, Mulu W, Genet C, Kibret M, Belete MA. Emergence of High Prevalence of Extended-Spectrum Beta-Lactamase and Carbapenemase-Producing Enterobacteriaceae Species among Patients in Northwestern Ethiopia Region. *Biomed Res Int* 2022**; 2022**: 5727638

106 Seni J, Najjuka CF, Kateete DP, et al. Antimicrobial resistance in hospitalized surgical patients: A silently emerging public health concern in Uganda. *BMC Res Notes* 2013; **6**: 298

107 Sewunet T, Asrat D, Woldeamanuel Y, et al. Polyclonal spread of blaCTX-M-15 through high-risk clones of Escherichia coli at a tertiary hospital in Ethiopia. *J Glob Antimicrob Resist* 2022; **29**: 405–12.

108 Soré S, Sanou S, Sawadogo Y, et al. Faecal carriage of extended-spectrum beta-lactamase-producing Enterobacteriaceae in healthy volunteers and hospitalized patients in Ouagadougou, Burkina Faso: prevalence, resistance profile, and associated risk factors. *African Journal of Clinical and Experimental Microbiology* 2021; **22**: 157–63.

109 Soré S, Sawadogo Y, Bonkoungou JI, et al. Detection, identification and characterization of extended-spectrum beta-lactamases producing Enterobacteriaceae in wastewater and salads marketed in Ouagadougou, Burkina Faso. *Int J Biol Chem Sci* 2020; **14**: 2746–57.

110 Teferi S, Sahlemariam Z, Mekonnen M, et al. Uropathogenic bacterial profile and antibiotic susceptibility pattern of isolates among gynecological cases admitted to Jimma Medical Center, South West Ethiopia. *Sci Rep* 2023; **13**: 7078.

111 Teklu DS, Negeri AA, Legese MH, Bedada TL, Woldemariam HK, Tullu KD. Extended-spectrum beta-lactamase production and multi-drug resistance among Enterobacteriaceae isolated in Addis Ababa, Ethiopia. *Antimicrob Resist Infect Control* 2019; **8**: 10.1186/s13756-019-0488-4.

112 Tellevik MG, Blomberg B, Kommedal Ø, Maselle SY, Langeland N, Moyo SJ. High prevalence of faecal carriage of esbl-producing enterobacteriaceae among children in Dar es Salaam, Tanzania. *PLoS One* 2016; **11**: e0168024

113 Tesfaye H, Alemayehu H, Desta AF, Eguale T. Antimicrobial susceptibility profile of selected Enterobacteriaceae in wastewater samples from health facilities, abattoir, downstream rivers and a WWTP in Addis Ababa, Ethiopia. *Antimicrob Resist Infect Control* 2019; **8**: 134

114 Tola MA, Abera NA, Gebeyehu YM, Dinku SF, Tullu KD. High prevalence of extended-spectrum betalactamase- producing Escherichia coli and *Klebsiella pneumoniae* fecal carriage among children under five years in Addis Ababa, Ethiopia. *PLoS One* 2021; **16**: e0258117.

115 Tufa TB, Mackenzie CR, Orth HM, Wienemann T, Nordmann T, Abdissa S, et al. Prevalence and characterization of antimicrobial resistance among gram-negative bacteria isolated from febrile hospitalized patients in central Ethiopia. *Antimicrob Resist Infect Control* 2022; **11**: 10.1186/s13756-022-01053-7

116 Tumuhamye J, Steinsland H, Bwanga F, et al. Vaginal colonization with antimicrobial-resistant bacteria among women in labor in central Uganda: prevalence and associated factors. *Antimicrob Resist Infect Control* 2021; **10**: 37

117 Yehouenou CL, Kpangon AA, Affolabi D, et al. Antimicrobial resistance in hospitalized surgical patients: a silently emerging public health concern in Benin. *Ann Clin Microbiol Antimicrob* 2020; **19**: 54

118 Zemtsa RJ, Noubom M, Founou LL, et al. Multidrug-Resistant and Extended-Spectrum β-Lactamase (ESBL)-Producing Enterobacterales Isolated from Carriage Samples among HIV Infected Women in Yaoundé, Cameroon. *Pathogens* 2022; **11**: 504
